# GPAS: an online AI system for rapid and accurate pathogen identification and LLM-based interpretation

**DOI:** 10.64898/2026.02.18.26346517

**Authors:** Tingting Li, Hao Hong, Duchangjiang Fan, Jin Li, Ting Li, Jiaqi Wu, Shuai Jiang, Xianxing Xie, Yawei Zhang, ManDong Hu, Xiaoyao Yin, Yizhe Zhang, Heping Ma, Zhehan Liu, Zhihui Su, Xiping Yu, Yu Liu, Hetian Yuan, Weifan Zheng, Haoyuan Liu, Mingyue Ma, Xingyue Li, Yezhuang Shen, Cheng Zhang, Yuyi Wang, Bing Zhao, Liming Sun, Qiuying Han, Jing Chen, Ke Zhang, Liang Chen, Na Wang, Weihua Li, Jianghong Man, Kun He, Fangting Dong, Fei Du, Yan Yi, Ailing Li, Tao Zhou, Xuemin Zhang, Tao Li

**Affiliations:** Nanhu Laboratory, State Key Laboratory of Biomedical Analysis (SKLBA, also known as National Center of Biomedical Analysis, NCBA), Beijing, China

## Abstract

Accurate identification of unknown pathogens is critical for medicine and public health, yet current metagenomic workflows remain heavily dependent on specialized bioinformatics expertise and manual interpretation, creating substantial bottlenecks in time-sensitive diagnostic settings^1^. The key challenges lie in achieving precise species identification amidst high background noise and translating complex microbial data into clinically actionable insights^2,3^. Here we present the Global Pathogen Analysis System (GPAS), an integrated computational framework that combines rapid and accurate pathogen identification with large language model (LLM)-based semantic interpretation. Central to GPAS is a dynamic-library alignment mechanism informed by prior probabilities of inter-species misclassification. By integrating a hybrid machine learning model that couples elastic neural networks with Bayesian inference, this approach substantially reduces both false positives and false negatives, achieving species-level accuracy superior to existing state-of-the-art tools. To enable clinical interpretation, we constructed a unified microbial knowledge graph integrating global metagenomic and metaviromic sample repositories, and trained a pathogen-specialized LLM agent. Through end-to-end reinforcement learning, the agent autonomously executes multi-step reasoning workflows extracting pathogen-specific insights from complex data and generating human-readable, evidence-based reports. Application to throat swab samples demonstrates that GPAS not only accurately identifies pathogenic microorganisms but also reveals how SLE-associated immune dysregulation reshapes the respiratory microbiome and promotes pathobiont overgrowth, providing clinically instructive interpretations. By substantially lowering technical barriers to pathogen identification, GPAS offers an accessible yet powerful platform for clinical diagnostics, public health surveillance, and microbiome research. The system is freely available at: https://gpas.nh.ac.cn/.

## Main Text

Rapid and accurate identification of unknown pathogens is fundamental to clinical diagnosis, public health surveillance, and outbreak response^1^. It is currently achieved primarily through metagenomic sequencing (mNGS), a powerful hypothesis-free approach, enables unbiased identification of all microorganisms^4,5^. However, broad implementation of mNGS in routine clinical practice remains constrained by several challenges^6^. First, analytical outputs are often accompanied by high background noise and substantial false positives. Second, complex lists of microbial taxa are difficult to translate into clinically actionable insights^3^. Third, interpretation relies heavily on highly trained bioinformaticians and microbiologists and is typically time-consuming, requiring expert judgement and multidisciplinary manual review.

A primary reason for these challenges is that different metagenomic classification tools, while each possessing their own advantages, struggle to simultaneously achieve both high sensitivity and high accuracy^7^. Current tools broadly follow two technical paradigms that embody distinct trade-offs. The first comprises exact k-mer matching methods^8^, represented by Kraken2^9^, which align read k-mers to reference genome databases and typically provide high sensitivity at species-level classification. However, when encountering highly homologous or conserved gene sequences, these methods are prone to misassigning reads among closely related species^10^. Moreover, the rapid expansion of metagenomic databases and increasing sequence redundancy have led to a growing number of highly conserved genomic regions shared across species, progressively diluting the discriminative power of characteristic k-mers^11^. A second class of methods, represented by Sylph^12^, adopts sketch-based strategies^13,14^ that compress reads and references into compact genomic fingerprints. These approaches offer high specificity but are less sensitive to low-abundance taxa.

Beyond accurate microbial identification, clinical interpretation of metagenomic data requires integrating multi-source information and knowledge-based reasoning—integrating host characteristics, sample provenance, disease phenotypes, and medical evidence to guide clinical decisions^2^. While this aligns with LLMs’ strengths in knowledge integration and logical inference^15^, no dedicated reasoning LLM agent exists for metagenomic interpretation^16^. Existing models like Evo^17^, LucaProt^18^ and DNABERT^19^ excel in sequence-level tasks but lack capacity to incorporate clinical semantics, microbial ecology, or pathogen-host causality. Developing LLMs with microbial knowledge reasoning capabilities—integrating metagenomic data with knowledge graphs, clinical literature, and multi-omics evidence—represents a critical frontier for advancing metagenomics from descriptive profiling toward mechanistic clinical insight^16,20-22^.

To address these challenges, we developed the Global Pathogen Analysis System (GPAS, https://gpas.nh.ac.cn), an integrated online platform for automated, high-precision pathogen identification and LLM-driven clinical interpretation (Fig. 1a). GPAS introduces three key innovations: a non-redundant, high-quality microbial genome database (GenoDB, Fig. 1b) constructed through similarity-based clustering to eliminate genomic redundancy while preserving comprehensive species coverage; a dynamic library alignment algorithm (Fig. 1c) that leverages pre-computed inter-species misclassification probabilities and a mixed statistical model to refine initial taxonomic assignments, drastically reducing both false positives and false negatives; a genome coverage pattern recognition framework that assesses the confidence of each identified species by comparing its coverage profile against reference distributions derived from NGS data of 24,164 samples. Building on this foundation, we developed GPAS-LLM, a specialized model fine-tuned on a curated pathogen knowledge graph encompassing 1,242 pathogen species, 10,493 comprehensively reviewed articles, and over 38.82M relational triplets. Operating within a multi-agent framework (Planner, Researcher, and Reflector), GPAS-LLM autonomously translates complex metagenomic data into evidence-based, clinically interpretable reports, providing a comprehensive solution for clinical and public health decision-making.

**Figure 1.**
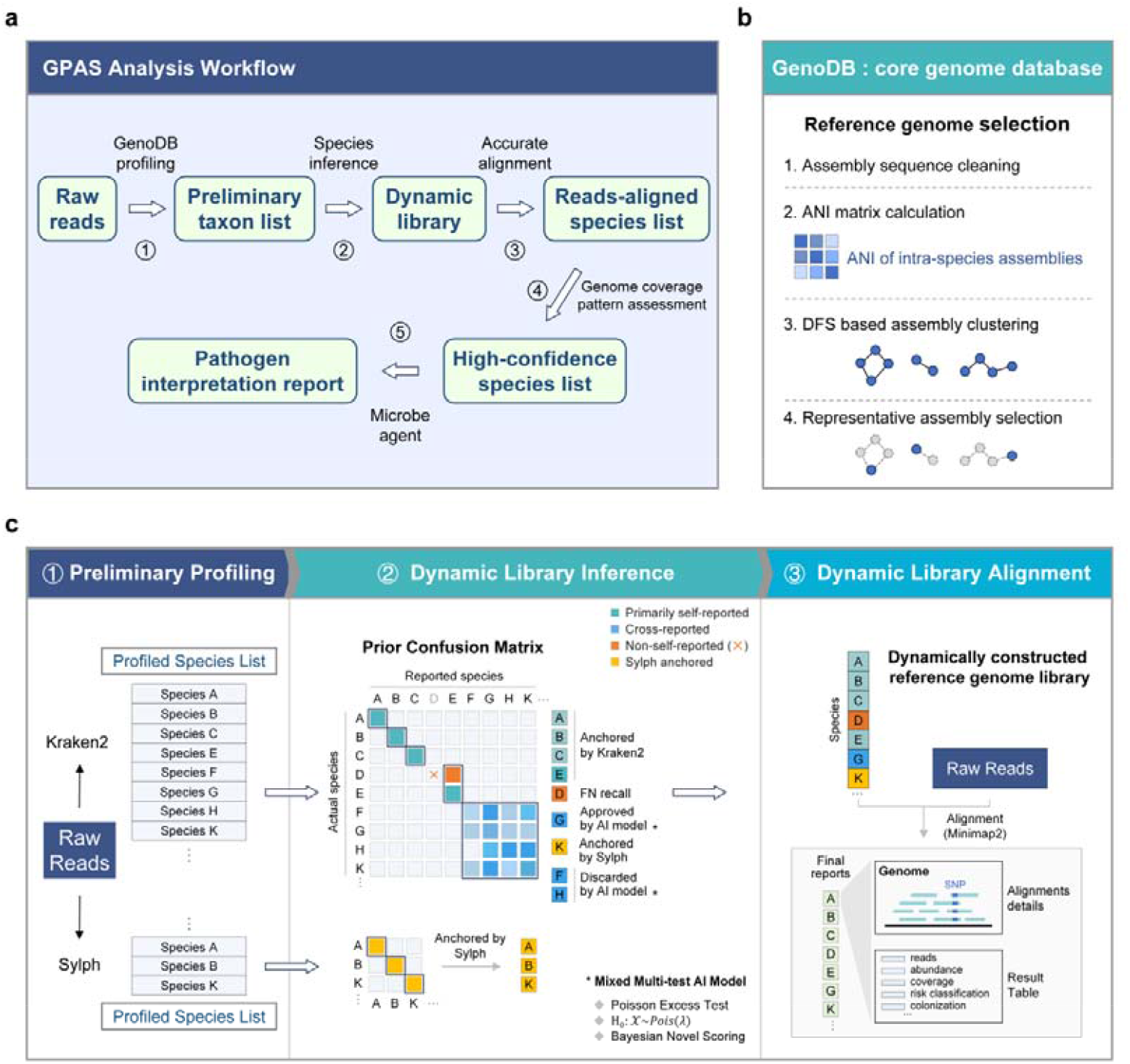
Overview of the Global Pathogen Analysis System (GPAS) framework. **a**, Schematic overview of the GPAS analysis workflow. Upon uploading raw sequencing data, the system sequentially performs preliminary profiling against the GenoDB database, dynamic library algorithm alignment, accurate alignment using dynamically constructed reference libraries, and genome coverage pattern-based confidence assessment, culminating in an intelligent report generated by the pathogen-specialized large language model (LLM) agent. **b**, Workflow for constructing the non-redundant genome database GenoDB. The pipeline encompasses genome sequence cleaning, intra-species average nucleotide identity (ANI) calculation, graph-based assembly clustering, and representative genome selection. **c**, Schematic illustration of the dynamic-library alignment algorithm. The algorithm integrates preliminary profiling results from Kraken2 (high sensitivity) and Sylph (high specificity) (c□), applies a hybrid statistical AI model calibrated by prior inter-species misclassification probability matrices (c□), and dynamically generates a high-confidence species list for subsequent validation by precise alignment using Minimap2 (c□).

### GenoDB: Non-redundant microbial genome database

A high-quality microbial reference genome database is essential for rapid and accurate species identification. Current reference database faces the dual challenges of rapid genome expansion and high sequence redundancy^23^. Extensive genome sequence similarity exists among different species, particularly within bacteria (Extended Data Fig. 1a–f), which not only compromises the accuracy and reliability of species identification but also limits computational efficiency for full-scale microbial data analysis^24^. To address these issues, we developed a representative genome selection strategy based on similarity clustering and constructed a de novo non-redundant genome database, GenoDB, covering all microbial species (Fig. 1b). By clustering intra-species genomes based on similarity and selecting a representative genome from each cluster, GenoDB reduced the overall database size to one-tenth of the original volume (Extended Data Fig. 2a–c). Compared with the Kraken2 default PlusPF database, GenoDB achieved broader species coverage, higher self-mapping rates, and lower unclassified read proportions (Extended Data Fig. 2d–g). Therefore, GenoDB enables full-spectrum microbial identification with enhanced accuracy and efficiency through systematic redundancy reduction.

### Dynamic library alignment algorithm (DLA)

To achieve species identification with both high accuracy and high sensitivity, we developed an innovative dynamic library alignment algorithm. The core principle of the algorithm is to calibrate the initial species lists generated by classifiers such as Kraken2 based on prior inter-species misclassification probabilities, dynamically generating a high-confidence species list (Fig. 1c). This algorithm comprises three steps: preliminary profiling, dynamic library inference, and dynamic library alignment (Fig. 1c). First, Kraken2 and Sylph were used jointly for preliminary species identification, leveraging the complementary advantages of the high sensitivity of the former and the high specificity of the latter to obtain a candidate species list. Subsequently, based on pre-constructed Kraken2 and Sylph prior misclassification matrices (Extended Data Fig. 3a–e and Extended Data Fig. 4a,b), a hybrid statistical AI model was introduced for comprehensive discrimination: low-confidence species were filtered out to control false positives, while potential species easily missed by conventional methods (i.e., false negatives) were recalled, thereby achieving a synergistic optimization of precision and sensitivity at the species level. Finally, based on the inferred species list, the system dynamically extracted corresponding reference genomes from the GenoDB database and performed sequence alignment using Minimap2^25^.

We trained and validated the DLA model on 40,000 simulated datasets comprising multi-species mixtures at varying sequencing depths (Extended Data Fig. 5a). The model used Sylph preliminary identification results as a basis, calibrating to select high-confidence species as anchor species. Results showed that the identification accuracy of anchor species reached as high as 0.995 (Extended Data Fig. 5b). The mixed prior abundance constructed based on anchor species showed good consistency with the raw abundance reported by Kraken2, with a mean normalized Jesen-Shannon divergence of 0.065 and variance of 0.153 (Extended Data Fig. 6a), indicating that the mixed prior distribution constructed by the model closely matched the measured data. Further analysis revealed that each anchor species could effectively eliminate an average of 50 false-positive species (Extended Data Fig. 6b).

### DLA performance evaluation

To comprehensively evaluate the species identification performance of GPAS, we systematically compared it with Centrifuger^26^, Kraken2 and Ganon2^27^ on multiple metagenomic datasets. The results showed that GPAS substantially reduced species misclassification rates (Fig. 2a–c). Evaluation of misclassification for all species at 10× sequencing depth revealed that while other tools exhibited at least 1-5 false positives in over 50% of species on average, GPAS achieved zero false positives for nearly all microbe species (99.8%) (Fig. 2a, Extended Data Fig. 7a). Across datasets at four different sequencing depths, GPAS consistently demonstrated extremely low false positives (Fig. 2b). Further analysis of inter-family misclassifications showed that the number of species incorrectly assigned to different families reported by GPAS was far lower than that of other tools (Fig. 2c). GPAS achieved higher species self-mapping rates (Fig. 2d). Evaluation of self-mapping rates at 10× depth showed that GPAS achieved near-perfect correct matches for the vast majority of species, while other tools generally exhibited higher unmapped proportions (Fig. 2d). In the dataset of 40,000 samples mixing different species and sequencing depths (Extended Data Fig. 5a), Kraken2 reported an average of 59.1 false positives per sample, whereas GPAS controlled false-positive species to an extremely low level, with an average of 0.7 (Fig. 2e).

**Figure 2.**
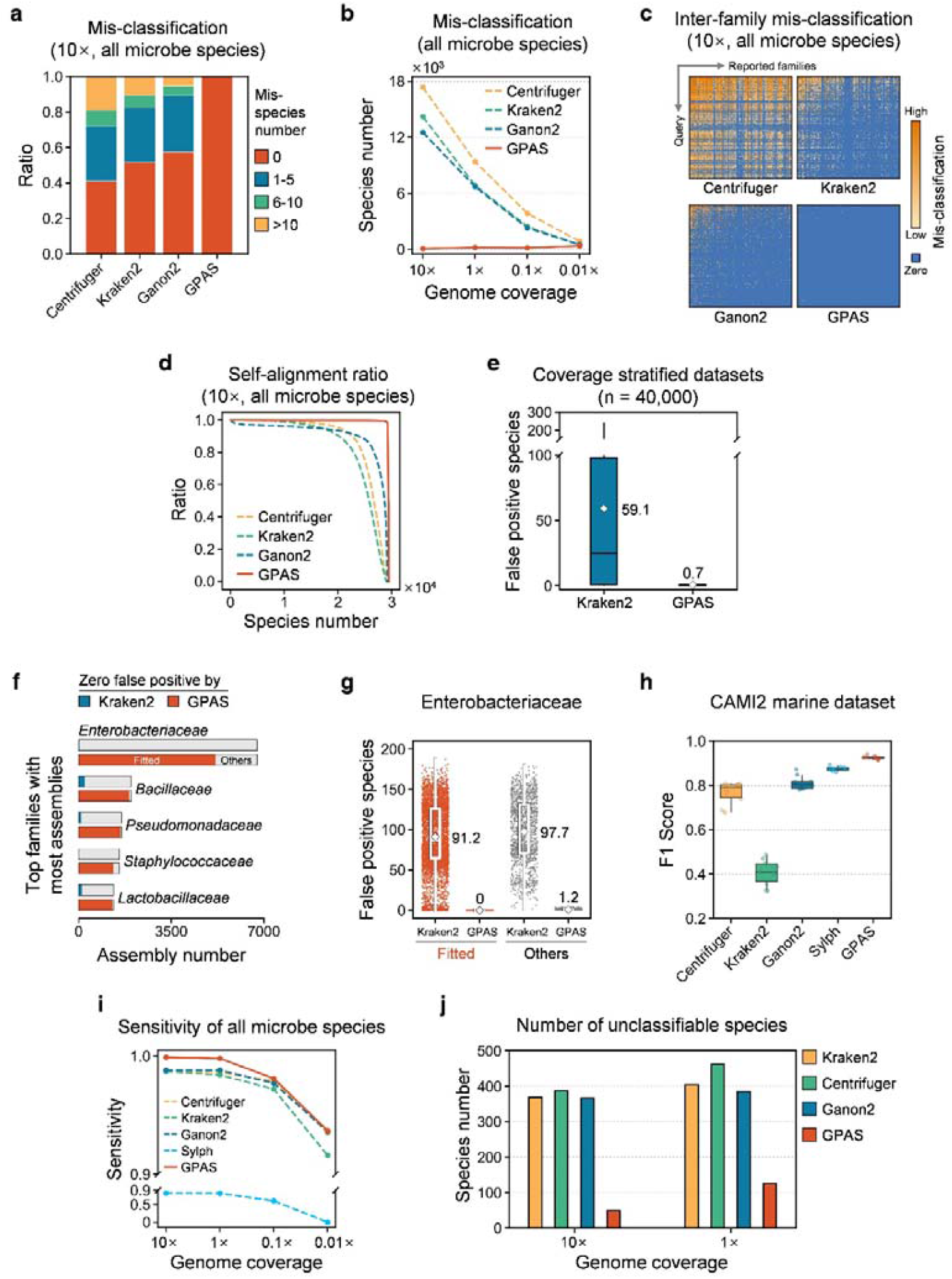
Comparative performance of GPAS across multiple datasets. **a**, Distribution of false positive species (supported by ≥5 reads) reported by four tools (Kraken2, Centrifuger, Ganon2, and GPAS) on simulated data at 10× genome coverage. GPAS achieves zero false positives for most species. **b**, Number of species with false positive reports (≥5 supporting reads) across four sequencing depths (10×, 1×, 0.1×, and 0.01×) for the four tools. GPAS maintains the lowest false positive counts at all depths. **c**, Inter-family misclassification events at 10× coverage (displaying the top 200 families with the highest species richness). GPAS reports substantially fewer cross-family misassignments compared to other tools. **d**, Species-level self-alignment ratio comparison among tools at 10× coverage. **e**, Average false positive species reported by Kraken2 and GPAS on a simulated training dataset comprising 40,000 multi-species mixtures. GPAS reduces the average false positives from 59.1 (Kraken2) to 0.7. **f**, Proportion of genomes achieving zero false positives for GPAS and Kraken2 within the five bacterial families with the highest assembly’s richness. Red: GPAS-fitted genomes (achieving zero false positives after GPAS hybrid model correction); blue: genomes achieving zero false positives with Kraken2. **g**, False positive reporting for all genomes within the family *Enterobacteriaceae* on 10× simulated data. The GPAS-fitted group (red) achieves zero false positives in 77.8% of samples, with remaining samples (grey) averaging 1.2 false positives, compared to Kraken2 averages of 91.2 and 97.7, respectively. **h**, Performance comparison (F1-score) of five tools on the CAMI II marine metagenome dataset. **i**, Species detection sensitivity comparison among five tools across four sequencing depths. GPAS maintains the highest sensitivity at all depths. **j**, Number of genomes that could not be correctly classified to their own species by four tools at 10× and 1× coverage. GPAS exhibits the fewest unclassified species, indicating the smallest detection blind spot.

Among the five bacterial families with the most species, GPAS achieved zero false positives in the vast majority of species identifications (Fitted), while Kraken2 achieved zero false positives for only a very small fraction of species (Fig. 2f). Within Enterobacteriaceae, a family characterized by high sequence similarity (Extended Data Fig. 1f), GPAS reduced false-positive species to zero in Fitted samples accounting for 76.6% of all assemblies, with an average of 1.2 false positives in the remaining assemblies, whereas Kraken2 exhibited average false positives of 91.2 and 97.7, respectively (Fig. 2g). On the CAMI II marine metagenome dataset^28^, GPAS achieved a F1-score of 0.925, substantially outperforming compared tools (Fig. 2h and Extended Data Fig. 7b). Furthermore, sensitivity analysis showed that GPAS maintained the highest sensitivity of species detection across datasets at four different sequencing depths (Fig. 2i). Meanwhile, GPAS exhibited the fewest unclassified species, indicating the smallest detection blackbox (Fig. 2j). Additionally, GPAS preloads the GenoDB database, thus achieves exceptional analysis speed (Extended Data Fig. 7c). In summary, GPAS effectively controls misclassification and false positives through dynamic library alignment algorithm, demonstrating excellent comprehensive identification performance.

### Genome coverage pattern recognition

GPAS innovatively introduced genome coverage pattern analysis for confidence assessment of species identification results. The core hypothesis of this method is that truly present species in metagenomic samples exhibit non-fragmented, uniquely patterned genome coverage, whereas false-positive identifications often arise from biased, fragmented genomic matches^29^. To validate this hypothesis, we used SARS-CoV-2 as an example, demonstrating its genome coverage patterns across 802 samples: regardless of high or low abundance, the positional distribution of sequences across the genome exhibited highly consistent coverage patterns (Fig. 3a). Based on t-SNE dimensionality reduction clustering analysis, different viral species displayed specific genome coverage patterns, and were clearly divided into distinct clusters, indicating that coverage patterns are species-specific (Fig. 3b). In comparison, most false-positive species detected across three real-world datasets exhibited highly biased, fragmented distributions (Extended Data Fig. 8a–e and Extended Data Fig. 9). Together, these findings demonstrate that true-positive and false-positive species show distinct differences in genome coverage, which can serve as effective features for species discrimination (Extended Data Fig. 10a–g).

**Figure 3.**
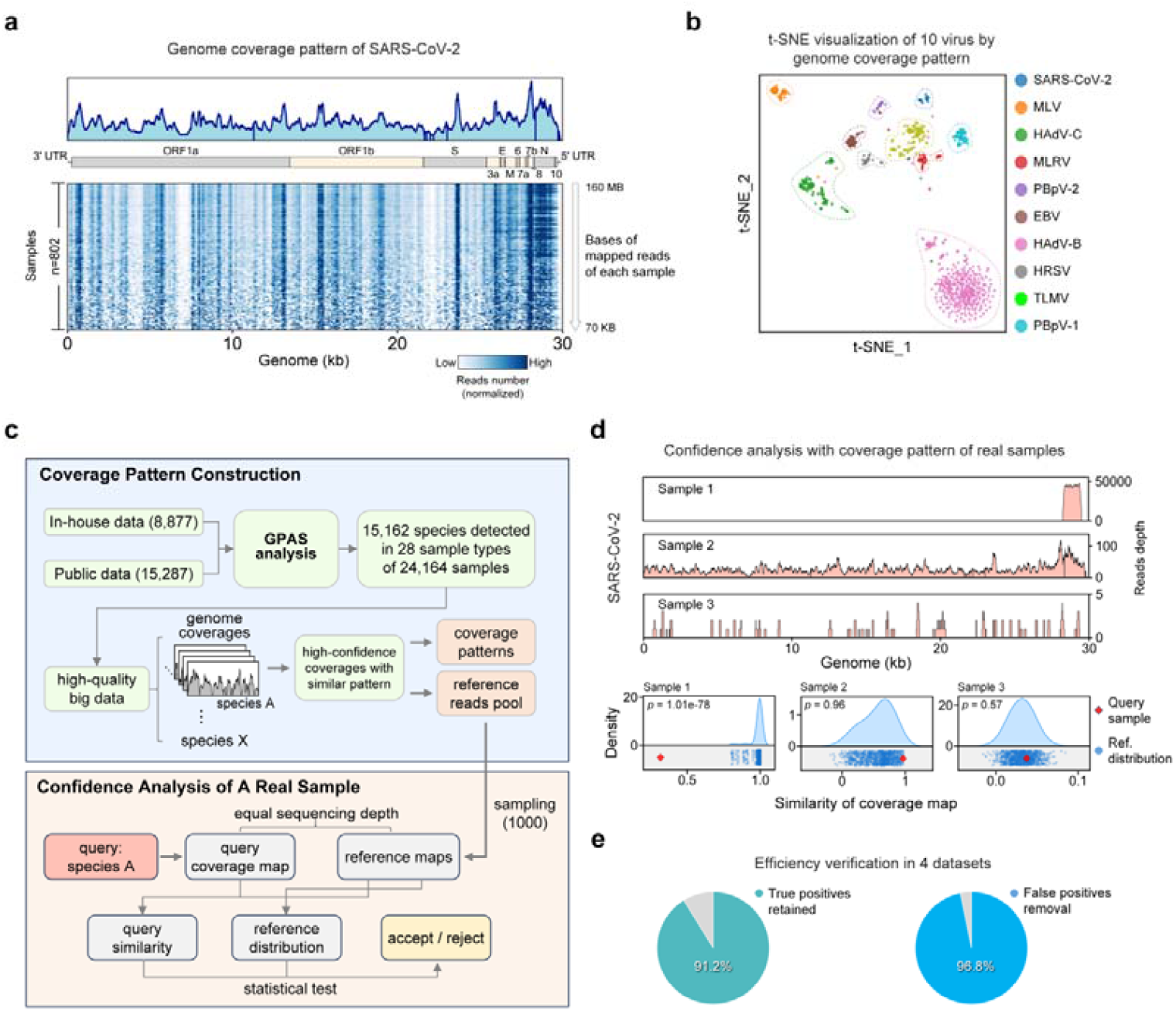
Genome coverage pattern recognition for reliability assessment. **a**, Genome coverage profiles of SARS-CoV-2 across 802 samples. Despite varying abundances, coverage patterns exhibit remarkable consistency across samples. **b**, t-SNE clustering of ten viruses based on genome coverage patterns. Viruses form distinct clusters according to coverage pattern similarity, demonstrating species-specific coverage signatures. **c**, Schematic of the statistical model for coverage pattern-based confidence assessment. A reference coverage library is constructed from 24,164 samples; confidence for query species is evaluated through equal-depth resampling and statistical comparison against reference distributions. **d**, Comparison of coverage patterns for three representative SARS-CoV-2 samples against reference distributions. Sample 1 shows significant deviation (p = 2.45e-60), suggesting false positive; samples 2 and 3, despite different sequencing depths, exhibit patterns highly concordant with references, supporting true positive detection. **e**, Validation results on four independent test datasets. The model retains 93.8% of true positive species while effectively removing 99.6% of false positives.

We established a statistical model for species genome coverage characteristics based on large-scale real metagenomic data (Fig. 3c). This model integrated 24,164 metagenomic samples covering 28 sample types, with a total of 15,162 microbial species detected (Extended Data Fig. 11). Based on these high-quality data, a high-confidence reference coverage pattern library was constructed for each species. For target species in test samples, the coverage profile was compared statistically with the reference distribution through equal-depth sampling to assess the reliability of identification results (Fig. 3c). Taking three representative SARS-CoV-2 samples as examples, the coverage profile of Sample 1 showed significant deviation from the reference distribution (p-value = 1.01e-78), suggesting potential false-positive identification; although Samples 2 and 3 differed substantially in sequencing depth, their coverage profiles were highly consistent with the reference distribution, with statistical tests supporting them as true positives (Fig. 3d). Validation on four independent test datasets (Extended Data Fig. 8a and Extended Data Fig.10c–g) demonstrated that the model effectively removed 96.8% of false-positive results while retaining 91.2% of true-positive species (Fig. 3e). These results indicate that confidence assessment based on genome coverage patterns can substantially improve species identification specificity without sacrificing sensitivity, providing a reliable quality control tool for clinical metagenomic interpretation.

### Pathogen intelligence agent

GPAS constructed a large language model-based pathogen interpretation agent. To achieve automatic transformation of metagenomic species identification results into clinically actionable insight, we designed a specialized large language model (GPAS-LLM) agent system for evidence-based pathogen interpretation (Fig. 4a). The system comprises two modules: first, a knowledge graph-based context engineering module that performs entity linking and standardization on the species list output by GPAS, retrieving relevant information from the constructed microbial knowledge graph to form an enhanced context window (Fig. 4a, left panel). This knowledge graph integrates all known pathogens of 1,242 species, 10,493 comprehensively reviewed articles, over 2.47 million microbial entities (six categories), and more than 38.82 million microbial relation triplets (nine categories, Extended Data Fig. 12a–e), incorporating 24,164 measured samples across 28 sample types (Extended Data Fig. 13a,b). Second, the pathogen interpretation agent architecture adopts a multi-agent collaboration mechanism: the Planner agent is responsible for user goal parsing and task decomposition, the Researcher agent invokes bioinformatics tools and the knowledge graph for evidence-based reasoning, and the Reflector agent performs error checking and feedback optimization, ultimately generating clinically evidence-based pathogen interpretation reports through GPAS-LLM (Fig. 4a, right panel). After model fine-tuning (Extended Data Fig. 14), GPAS-LLM was systematically compared with the baseline model (Qwen3-Next-80B-A3B-Instruct) on four authoritative biomedical benchmark datasets. Results showed that the fine-tuned model exhibited systematic enhancement in key dimensions including pathogen identification, clinical reasoning, and literature understanding, demonstrating robust performance improvement (Fig. 4b,c).

**Figure 4.**
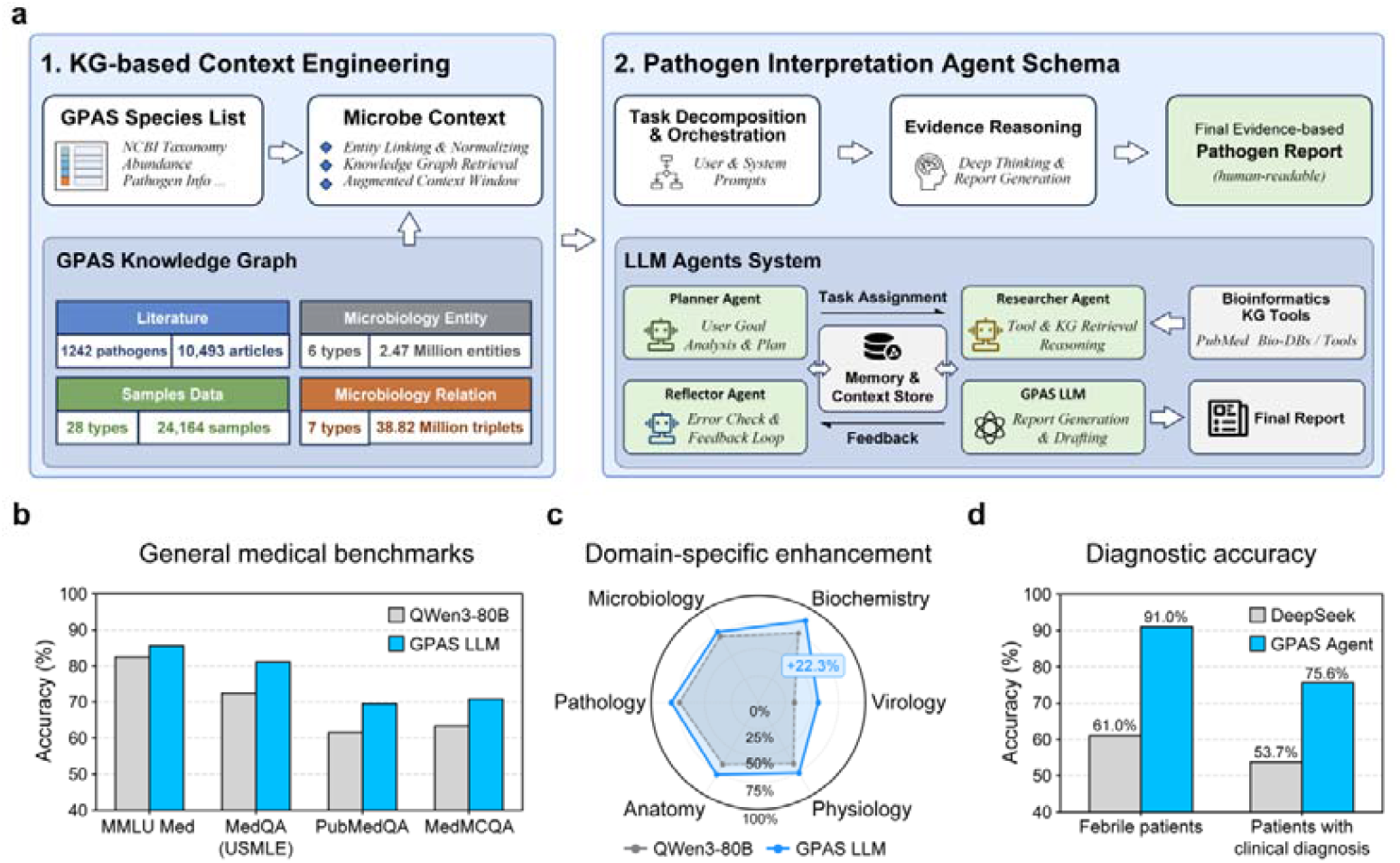
A metagenomic specialized LLM agent for evidence-based pathogen interpretation. **a**, Architecture of the GPAS-LLM agent system (right panel) and knowledge graph context engineering module (left panel). The Planner agent parses user goals and decomposes tasks; the Researcher agent performs evidence-based retrieval using integrated bioinformatics tools and knowledge bases; the Reflector agent executes error checking and feedback optimization; the GPAS-LLM generates final clinical interpretation reports. The knowledge graph integrates 1,242 pathogenic species, 10,493 curated articles, over 2.47M entity nodes, and 38.82M relation triplets. **b**,**c**, Performance comparison between GPAS-LLM and baseline model (Qwen3-80B) on four biomedical benchmarks. Fine-tuned models exhibit substantial enhancement in pathogen recognition, clinical reasoning, and literature comprehension. **d**, Clinical validation on real-world samples. Left: among 100 febrile patient throat swab samples, GPAS-LLM accurately interpreted fever-related symptoms for 91.0% of samples, significantly outperforming DeepSeek V3.2 (61.0%). Right: across 82 clinically diagnosed samples from diverse specimen types, GPAS-LLM achieved 75.6% accuracy in causative pathogen identification, compared to 53.7% for DeepSeek V3.2.

To validate the system’s efficacy in real clinical scenarios, we compared the GPAS agent with DeepSeek^30^ V3.2 using real metagenomic sequencing data, focusing on evaluating accuracy in clinical reasoning and diagnostic judgment. Analysis of 100 oropharyngeal swab metagenomic datasets from febrile patients showed that the GPAS agent accurately interpreted fever and related clinical symptoms in 91.0% of samples, significantly higher than DeepSeek V3.2’s 61.0% (Fig. 4d, left panel; Extended Data Fig. 15a). In another cohort of 82 samples with confirmed clinical diagnoses (covering sample types of respiratory infections including sputum, blood, lung tissue, and bronchoalveolar lavage fluid), the GPAS agent identified pathogenic microorganisms with 75.6% accuracy, outperforming DeepSeek V3.2’s 53.7% (Fig. 4d, right panel; Extended Data Fig. 15b). These results demonstrate that through deep integration of multi-agent collaboration and knowledge graphs, the GPAS agent exhibits clinical reasoning capabilities surpassing general-purpose large language models in complex infection cases.

### Application of GPAS in oropharyngeal swab sample analysis

To validate the practical application efficacy of GPAS, we performed deep sequencing on an oropharyngeal swab sample from a febrile patient with systemic lupus erythematosus (SLE) and conducted species identification using both Kraken2 and GPAS. Results showed that GPAS identified 201 microbial species in this sample, representing a >90% reduction compared to the 2,345 species reported by Kraken2, demonstrating GPAS’s significant advantage in controlling false positives (Fig. 5a). Further analysis of microbial community characteristics revealed that both the number of detected species and alpha diversity in this sample were significantly higher than those in oropharyngeal swab samples from healthy controls (Fig. 5b), suggesting unique microecological features in this SLE patient sample. Large-scale data-based sample type clustering analysis also accurately classified this sample as originating from oropharyngeal swabs (Fig. 5c), indicating that GPAS preserves the core microbiome structural characteristics^31^ while filtering false-positive identifications. At the output level, the GPAS system generates structured microbial identification lists, providing colonization status, risk level, confidence radar charts, genome coverage maps, and coverage pattern-based statistical confidence test results for each detected microorganism (Fig. 5d–f). Taking Neisseria subflava as an example, its genome coverage reached 85.3%, with coverage patterns highly consistent with the reference distribution (p = 0.51), and the confidence radar chart comprehensively evaluated its identification credibility across multiple dimensions including species confusion, abundance, and pattern recognition (Fig. 5e). Further analysis of assembled *N. subflava* sequences for virulence factors and drug resistance genes identified glycopeptide resistance-associated genes and multiple virulence factors, providing molecular-level evidence for understanding its potential pathogenic mechanisms (Fig. 5f). Based on this, the GPAS agent automatically parses the species identification list and generates an integrated pathogen analysis report (Fig. 5g). In this case, the agent accurately recognized that the microbial community in the sample primarily consisted of typical commensals or pathobionts of the oral cavity and upper respiratory tract, while noting that the abundances of multiple genera significantly deviated from normal ranges, strongly suggesting severe dysbiosis or acute infection status. Health status assessment further indicated that elevated microbial diversity implies impaired host immune function, which may interact bidirectionally with SLE-related immune dysregulation or febrile status^32^, collectively contributing to susceptibility to respiratory infection. In summary, through precise species identification and multi-dimensional confidence assessment, GPAS achieves deep interpretation from microbial composition analysis to clinical significance in oropharyngeal swab samples, providing an end-to-end solution for the clinical application of metagenomic data in real-world samples.

**Figure 5.**
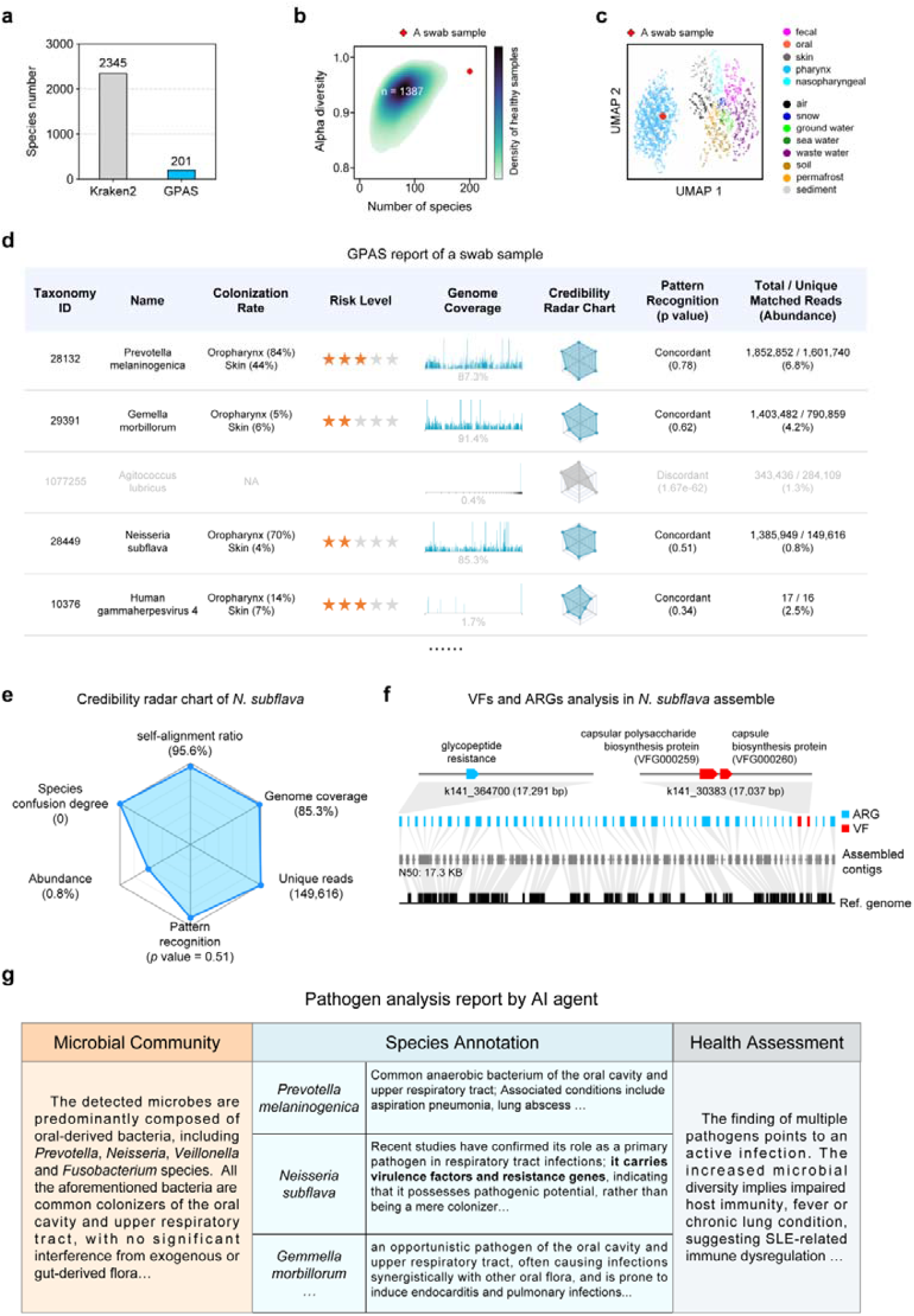
GPAS: from microbial identification to clinical insight in a swab analysis. **a**, Comparison of species detected by GPAS versus Kraken2. GPAS identified 201 microbial species, representing a >90% reduction compared to Kraken2 (2,345 species). **b**, Alpha diversity comparison between the SLE patient sample and healthy control oropharyngeal swabs (n = 1,387). The SLE patient sample exhibits significantly elevated species richness. **c**, UMAP clustering of samples based on taxonomic profiles. The SLE patient sample is accurately classified within the pharynx cluster. **d**, Representative structured output from the GPAS microbial identification interface, displaying multi-dimensional annotations for detected species. **e**, Confidence assessment details for *N. subflava*: genome coverage 85.3%, coverage pattern highly concordant with reference distribution (p = 0.51); radar chart comprehensively visualizes multi-dimensional confidence metrics including species confusion degree, abundance, and pattern recognition score. **f**, Virulence factor and antimicrobial resistance gene analysis of assembled *N. subflava* sequences, identifying glycopeptide resistance genes and multiple virulence factors. **g**, Pathogen analysis report generated by the GPAS-LLM agent, encompassing microbial community summary, species-level annotations with clinical implications, and integrated health assessment.

## Discussion

The Global Pathogen Analysis System (GPAS) presented in this study addresses two fundamental challenges that have long constrained the clinical application of metagenomic sequencing: the technical difficulty of accurate species identification in complex microbial backgrounds, and the interpretive gap between sequence data and clinically meaningful insights. By integrating a dynamic-library alignment mechanism with a large language ^33^model-powered reasoning agent, GPAS establishes an end-to-end computational framework that transforms raw sequencing data into clinical actionable insights with minimal human intervention.

While public repositories such as RefSeq offer unprecedented breadth of microbial genome sequences, their growing redundancy has emerged as a primary source of false-positive identifications. The clustering-based, redundancy-minimized GenoDB database we constructed addresses this bottleneck by preserving species-specific genomic signatures while reducing database size by an order of magnitude. More importantly, the dynamic-library alignment mechanism moves beyond static database matching by intelligent species inferencing informed by inter-species misclassification priors. This design enables GPAS to achieve what individual tools cannot: simultaneously harnessing the sensitivity of k-mer-based methods and the precision of alignment-based approaches while suppressing their respective limitations. The substantial reduction in false positives from thousands of species to hundreds in clinical samples underscores the practical impact of this strategy for real-world diagnostics.

Beyond taxonomic classification, this study introduces a conceptually novel approach to result validation through genome coverage pattern analysis. The observation that true-positive species exhibit consistent, non-fragmented coverage profiles across samples, while false positives arise from sporadic, random matches, provides a statistically grounded basis for confidence assessment. By constructing species-specific reference coverage distributions from over 24,000 samples, GPAS enables quantitative evaluation of individual detection events. This coverage-based validation framework offers a complementary dimension to traditional abundance- or uniqueness-based metrics.

The clinical utility of GPAS is perhaps most vividly illustrated in its application to SLE patients with fever. Here, the system identified not only oral commensals but also elevated microbial diversity and pathobiont overgrowth. Through the LLM agent, these findings pointed to underlying immune dysregulation as a predisposing factor for opportunistic infection. This capacity to link microbial community features with host pathophysiology exemplifies the paradigm shift from descriptive microbiome profiling to mechanistic clinical insight. By revealing how SLE-associated immune dysfunction reshapes the respiratory microbiome and promotes pathobiont expansion, GPAS offers a window into host-microbe interactions that static species lists cannot provide.

Despite these advances, several limitations warrant consideration. First, while GenoDB substantially reduces redundancy, its coverage of understudied or recently discovered pathogens remains dependent on the underlying RefSeq database. Continuous updating mechanisms will be essential to maintain relevance as novel pathogens emerge. Second, the coverage pattern validation framework, though robust for well-characterized species, may encounter limitations for extremely rare or poorly sampled organisms where reference distributions cannot be reliably established. Third, while the LLM agent demonstrates strong performance on curated test sets, its real-world clinical deployment will require rigorous prospective validation across diverse populations, sample types, and disease contexts. The potential for model hallucinations though mitigated by knowledge graph grounding and multi-agent reflection cannot be entirely eliminated and warrants ongoing scrutiny.

Looking forward, several directions merit exploration. Integration of longitudinal sampling could enable GPAS to track within-host microbial dynamics over disease courses and treatment responses, offering prognostic as well as diagnostic value. Expansion of the knowledge graph to incorporate metabolomic, immunologic, and host genetic data could further enrich interpretive depth, moving toward multi-omic integration. From a translational perspective, prospective clinical trials comparing GPAS-assisted diagnosis with conventional workflows are needed to quantify its impact on clinical outcomes such as time to appropriate therapy, antibiotic stewardship, and patient survival. Additionally, as antimicrobial resistance continues to escalate as a global health threat^34^, extending the framework to provide real-time resistance gene profiling and mechanistic interpretation could substantially enhance its clinical impact.

In conclusion, GPAS represents a comprehensive computational ecosystem that redefines the boundaries of metagenomic diagnostics. By harmonizing algorithmic innovation with AI-driven semantic interpretation, it bridges the longstanding gap between sequencing capacity and clinical applicability. As infectious diseases continue to challenge global health systems, platforms that democratize access to advanced pathogen analysis while preserving analytical rigor will play an increasingly pivotal role. We envision GPAS not merely as a tool, but as a foundational infrastructure for a future where “pathogen-first” precision diagnostics become integral to routine clinical practice.

## Data Availability

All data produced in the present study are available upon reasonable request to the authors

## Methods

### GenoDB database construction

The GenoDB database encompasses four microbial domains—archaea, bacteria, fungi, and viruses—along with eukaryotic host genomes. The construction workflow began with data acquisition from the NCBI RefSeq database (15 June 2025), followed by multi-stage quality control filtering to generate a high-quality, low-redundancy reference genome collection. Assembly summary tables and corresponding genomic sequences (*\*_genomic.fna.gz*) and annotation files (GFF/GTF formats) were downloaded for each domain. Concurrently, the NCBI Taxonomy database (*taxdump.tar.gz*) was downloaded, and the taxonomy of each assembly was updated using the taxdumpy tool (https://pypi.org/project/taxdumpy/) to replace merged or obsolete taxonomy IDs (taxids) with current valid taxonomic identifiers.

### Plasmid sequence removal

For prokaryotic genomes (archaea and bacteria), plasmid reference sequences were downloaded from the NCBI RefSeq plasmid database. After indexing with samtools^35^, plasmid accession identifiers were cross-referenced against contigs in each assembly. A contig was removed if its accession number appeared in the plasmid reference set or if its sequence description contained the keyword “plasmid”. Assemblies for which post-filtering total length fell below 40% of the original genome size were retained in their original form to avoid inadvertent deletion.

### Genome tiered filtering

For each species (taxids with rank = species), assemblies were prioritized according to NCBI assembly level (Complete Genome > Chromosome > Scaffold > Contig), retaining the highest-level assemblies for each species. For eukaryotic host species, only a single manually curated reference genome was retained per species. Assemblies with species names containing ambiguous or provisional designations such as “sp.”, “candidatus”, “unidentified”, or “uncultured” were excluded. For major eukaryotic host domains (invertebrates, plants, mammals, other vertebrates), only species with Complete Genome or Chromosome level assemblies were retained to ensure reference sequence quality.

### Average nucleotide identity calculation and clustering

For multiple assemblies within each species, pairwise average nucleotide identity (ANI) was computed. ANI for prokaryotes and fungi was calculated using fastANI^24^ and skani^36^; for viral genomes, given their high sequence diversity, lz-ani^37^ was employed with kmer-db (k=25) pre-filtering to improve computational efficiency.

### Representative genome selection

Based on ANI results, a genome similarity graph was constructed using a 95% ANI threshold. Initial clusters were identified through connected component analysis; loosely connected clusters with internal density below 0.8 were further subdivided using the Louvain community detection algorithm. Within each cluster, the assembly with the highest N50 value (or longest total length) was selected as the representative genome.

### Kraken2 database compilation

The final set of representative genomes was used to build a Kraken2 classification database. Genome sequences underwent low-complexity region masking with dustmasker^38^ before Kraken2 indexing (default parameters: k-mer length 35, minimizer length 31, spaced mask 7) using NCBI taxdump. For large eukaryotic genomes, an in-house Rust tool (*genome_reduce*), was developed to extract gene regions with flanking sequences based on GFF annotations, compressing genomes to functional regions to reduce database size.

### Simulation of sequencing reads

To establish empirical prior distributions for classifiers, simulated sequencing was performed for all microbial genomes (archaea, bacteria, fungi, viruses) in the GenoDB reference collection. Simulations were conducted using the NGSNGS^39^ tool, generating paired-end Illumina reads of 150 bp length for each assembly. Circular genome mode (-circ) was enabled for prokaryotic genomes to mimic authentic circular chromosome sequencing characteristics. Base substitution quality distributions were introduced based on empirical Illumina quality frequency files (AccFreqL150R1/R2), together with insertion and deletion errors (insertion and deletion rates both 0.1%, length parameter 0.1). A deterministic random seed derived from the assembly accession hash value was used for each assembly to ensure reproducibility.

Simulations were performed at two coverage depths: high coverage (10×) and medium coverage (1×). Low coverage (0.1×) and ultra-low coverage (0.01×) datasets were generated by down-sampling the 1× coverage data using seqtk, enabling evaluation of classifier performance across varying sequencing depths. The resulting FASTQ files were processed by two classification tools: (1) Kraken2, using the GenoDB custom database for k-mer classification with confidence threshold 0.1, outputting standard Kraken2 report format; (2) Sylph, a k-mer sketch-based metagenomic profiling tool, outputting species abundance profiles.

### Prior confusion matrix construction

Empirical prior databases were constructed separately for Kraken2 and Sylph based on the simulated classification results, serving as calibration references for the dynamic library algorithm. For the Kraken2 prior, classification reports for each assembly were parsed to extract species-level read counts (clade count). For each true species (true_taxid) *i*, the number of reads classified to each reported species (reported_taxid) *j* was recorded as *n*_*ij*_, constructing confusion records. Confusion records across all assemblies were aggregated by species pair (*i, j*) to calculate confusion probability: *P*(*j* | *i*) = ∑ *n*_*ij*_/ ∑ *N*_*i*_, where *N*_*i*_ represents the total simulated reads for species *i* across all assemblies. The resulting species-level aggregated confusion matrix was filtered to remove sparse entries with insufficient total read counts. Additionally, lineage profiles spanning species to class taxonomic ranks were extracted, recording clade counts and direct counts at each level for hierarchical validation.

For the Sylph prior, classification results across the three coverage gradients were processed to construct both forward and inverse aggregation profiles. Forward aggregation recorded which assemblies were reported by Sylph for each simulated assembly along with their abundances, enabling assessment of self-reporting rates and cross-reporting patterns. Inverse aggregation, from the perspective of reported assemblies, tallied which simulated sources could generate each report, calculating self-reporting probability *P*(true = *X*|report = *X*). Based on inverse aggregation results, each assembly was assigned an inverse reliability category for each coverage depth: HIGH (exclusively self-reporting), MEDIUM (self-reporting probability ≥50%), LOW (self-reporting probability <50%), ALIAS (deterministic redirection to another assembly), and FP_MAGNET (false positive attractor, reportable from multiple non-self sources). This reliability classification directly informed anchor selection in the dynamic library algorithm to distinguish high-confidence detections from potential false positive signals.

### Dynamic library alignment algorithm

GPAS implements a probabilistic framework for metagenomic species detection that integrates the high-specificity signals from Sylph alignment with the high-sensitivity signals from Kraken2 classification. The core strategy involves three steps: screening high-confidence anchor species from Sylph results, constructing a hybrid prior model to explain observed Kraken2 classification signals, and subsequently identifying potential novel species from residual signals.

#### Anchor species selection

For each genome *a* appearing in the Sylph alignment report, effective coverage was first used to assign it to coverage bin *b*. Based on Sylph prior reliability categories *r*_*d*_ across different simulated depths, each reliability category received a predefined score *score*(*r*_*d*_) ∈ [-1,1]. A weighted composite score was calculated for each genome *a*:

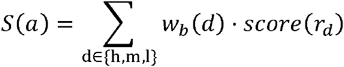

where *w*_*b*_(*d*) represents the weight assigned to depth *d* for coverage bin *b*. Genomes with *S*(*a*) ≥ *τ*_*anchor*_ were designated as anchor genomes, and their corresponding species were added to the anchor species set 𝒜.

#### Hybrid prior construction

Given anchor genomes {*a*_1_, *a*_2_,…, *a*_*K*_} with corresponding sequence abundance weights {*w*_1_, *w*_2_,…, *w*_*K*_}, normalized weights were calculated as 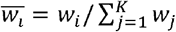 Using the Kraken2 prior confusion matrix, species-level confusion patterns *P*(*s*|*a*_*i*_) were obtained for each anchor genome *a*_*i*_. The species-level hybrid prior was defined as:

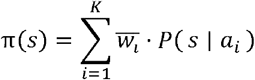

To evaluate the deviation between observed and expected counts for non-anchor species in the hybrid prior, an overdispersion factor was estimated using Pearson residuals across the hybrid prior sample set 𝒮:

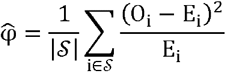

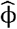 was then truncated to the interval [1.0, 5.0]. One-sided excess tests were performed for each candidate species in the hybrid prior: when 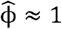, the Poisson exact test was applied for expected counts *λ*< 20, otherwise normal approximation with continuity correction was used; when *λ*< 20, a negative binomial test was employed with variance set as 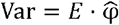, yielding test statistic 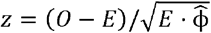. The resulting p-values were Benjamini-Hochberg corrected to obtain *q-*values; species with *q* < *α* (default *α* = 0.05) were classified as true positives.

#### Genus-level deconvolution and hierarchical validation

Building upon the hybrid prior, a residual classification tree was derived from the original Kraken2 classification tree using Euler tour-encoded difference arrays, achieving *O*(*n* + *m*) complexity (*n* = number of nodes, *m* = number of signals). For candidate species not included in anchor set 𝒜 nor appearing in hybrid prior *π*, yet retaining signals in the residual classification tree, an iterative deconvolution strategy was employed. Upon acceptance of a species *S*^*^, the false positive reads it contributed to other species *S*^′^ within the same group were estimated using the prior confusion matrix:

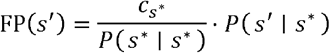

Estimated false positives were subtracted from read counts of remaining candidate species, and iteration continued until no candidate species met domain-specific thresholds.

### Big data collection and analysis

A total of 24,164 metagenomic and metaviromic sequencing datasets were included in this study. These included 7,716 publicly available metagenomic datasets collected from our previously established MAGdb^40^ resource and 7,571 publicly reported virome datasets curated from high-quality peer-reviewed studies. In addition, 8,877 in-house samples were collected and sequenced by NGS. Samples from participants were collected in accordance with medical ethics requirements, under the approval of the Ethics Committee of National Center of Biomedical Analysis (NCBA, Approval No. AF/SC-08/02.240N and AF/SC-08/02.453N). Written informed consent was obtained from all participants. All datasets were processed and analyzed using the GPAS framework under a unified analytical pipeline to ensure methodological consistency. Microbial detection was considered positive only when the genome coverage exceeded 5%.

### Confidence assessment based on genome coverage patterns

To establish a robust framework for detection confidence assessment, we developed a genome coverage pattern–based statistical validation strategy. For any detected species whose genome assembly was observed in ≥10 independent samples, a species-specific coverage feature profile was constructed. The top 50% of samples ranked by genome coverage were selected as seed samples and incorporated into an initial reference sample set, as these samples were assumed to provide the most reliable and complete genomic representation. Reads from these samples were pooled to form a reference reads pool. Coverage maps with sequencing depth ≥5× were designated as reference maps. For the remaining 50% of samples, coverage pattern consistency was evaluated against the reference sample set using statistical testing. Samples passing the statistical threshold were iteratively incorporated into the reference set. This procedure was repeated up to five rounds to obtain the final reference sample set, reference reads pool, and reference maps.

For confidence assessment of a query sample, two scenarios were considered:

1. Sequencing depth ≥5×:

The coverage map of the query sample was directly compared with all reference maps. The maximum similarity was extracted and evaluated against the similarity distribution among reference maps.

2. Sequencing depth <5×:

Given the potential incompleteness of genome coverage at low depth, theoretical coverage maps were simulated. Reads were randomly sampled from the reference reads pool to match the query sequencing depth, generating 1,000 simulated coverage maps. The mean pairwise similarity among simulated maps was calculated to construct a reference similarity distribution. The mean similarity between the query sample and all simulated maps was then statistically evaluated against this distribution.

Similarity between coverage maps was quantified using two complementary metrics:

- Jaccard similarity, measuring the overlap of covered genomic regions
- Pearson’s correlation coefficient (R), measuring similarity of coverage depth patterns

Statistical evaluation was performed using a Z-test. A detection was considered credible only when both similarity metrics passed the statistical threshold.

### Benchmarking among taxonomic classification tools

GPAS was benchmarked against Centrifuger, Kraken2, and Ganon2 using both the CAMI2 benchmark dataset and custom-built simulated datasets generated at varying sequencing depths to assess robustness under different abundance scenarios.

All tools were evaluated using the same reference database (GenoDB) to eliminate database-related bias. Default parameters recommended by each tool were used unless otherwise specified. Performance metrics included sensitivity, precision, F1-score, abundance estimation accuracy, and computational efficiency. Evaluations were performed across multiple sequencing depth conditions to comprehensively characterize classifier performance.

### Clinical dataset for intelligent agent performance evaluation

Two independent mNGS datasets were used to evaluate the performance of the GPAS intelligent agent and DeepSeek in report interpretation and pathogen identification. The first dataset comprised 100 samples from febrile patients and was used to assess the accuracy of report interpretation. The second dataset consisted of 82 samples with laboratory-confirmed pathogens, encompassing six types of upper respiratory tract specimens. This dataset was used to evaluate pathogen identification accuracy, defined as the ability of the intelligent agents to correctly identify the causative microbial species consistent with laboratory-confirmed diagnoses.

Both the GPAS intelligent agent and DeepSeek were evaluated using identical datasets and input conditions to ensure a fair and unbiased comparison. Performance was quantified based on report interpretation accuracy and pathogen identification accuracy relative to the established ground truth.

### Pathogen knowledge graph construction

A set of 1,242 pathogenic species with well-established clinical significance was curated based on authoritative disease databases, including the “Catalogue of Pathogenic Microorganisms Transmissible among Humans” (National Health Commission of the People’s Republic of China, 2023) and the NIAID Emerging Infectious Diseases Pathogens list, supplemented by clinical case reports. PubMed literature searches were conducted for these pathogens, restricted to the past decade (2016–2026) and high-impact factor journals. Retrieved abstracts were initially screened using the DeepSeek-V3.2 model^30^ to identify literature containing key information on pathogenic virulence mechanisms, pathogenicity, and toxicity characteristics. Preliminary screening results underwent manual verification to exclude false positives. For pathogens lacking relevant literature, search scopes were expanded and re-verified. The resulting high-quality literature dataset (n = 10,493 articles) was used for supervised fine-tuning of large language models.

### Large language model fine-tuning

A context-aware framework was developed for automated biomedical knowledge graph construction, comprising three phases: self-evolving open information extraction, knowledge fusion with semantic alignment, and agent-based collaborative orchestration.

The core data structure adopted a composite triplet schema: (Subject: [Attribute], Relation: [Condition], Object: [Attribute]), designed to capture fine-grained entity attributes (e.g., gene mutation sites) and relational conditions (e.g., experimental contexts) from biomedical literature.

### Self-evolving open information extraction

To address the scarcity of domain-specific annotated data, an iterative self-evolving learning strategy was implemented. Initial model training utilized a cold-start dataset comprising 1,000 core articles manually annotated by biomedical domain experts for key entities, relationships, and themes.

#### 1. Seed model construction

A seed model based on the Qwen3-8B architecture was supervised fine-tuned on the core literature dataset, equipping it with preliminary composite triple extraction capabilities.

#### 2. Pseudo-label generation and collaborative filtering

The model performed inference on large-scale unannotated literature and textbook texts to generate candidate composite triples. A collaborative filtering module with dual-threshold mechanism was subsequently applied for quality control: samples were retained as high-quality pseudo-labeled data only when prediction confidence exceeded *τ*_*conf*_ = 0.8 and information entropy fell below *τ*_*ent*_ = 0.3; samples failing these thresholds were discarded.

#### 3. Iterative fine-tuning

High-quality data retained after filtering were fed back into the training set for subsequent rounds of iterative model fine-tuning, progressively enhancing model generalization in open-domain scenarios.

### Knowledge fusion and alignment

To resolve semantic ambiguity and achieve standardization in extracted content, dual-path alignment processing was applied to raw composite triples for both entities and relations.

#### 1. Entity normalization

The SapBERT encoder generated semantic vectors for subjects and objects, computing similarity scores against the UMLS (Unified Medical Language System) concept repository. Entities with similarity scores exceeding the preset threshold (0.8) were normalized to existing UMLS concepts; those falling below threshold were incorporated as novel entities to preserve emerging terminology from frontier literature.

#### 2. Relation alignment

The BGE-m3 encoder vectorized relation descriptions for matching against standard relations in the UMLS semantic network. Using the same 0.8 threshold, relations were either mapped to standard relation types or dynamically extended as new relation patterns.

Following these processes, the system output standardized context-aware triples, ensuring semantic consistency throughout the knowledge graph.

### Agent-based collaborative orchestration

The entire construction and application workflow was coordinated by an Agent Controller based on the Model Context Protocol (MCP), enabling modular multi-agent collaboration.

#### 1. Multi-agent collaborative construction

The Extraction Agent performed raw information extraction from texts; the Normalization Agent executed the semantic alignment logic; the Graph Construction Agent ultimately wrote the curated structured data into the Neo4j graph database, instantiating the context-aware knowledge graph.

#### 2. Application interface

The QA Agent interacted with the underlying graph through a retrieval-augmented generation (RAG) interface, supporting downstream complex biomedical question-answering tasks.

## Data availability

The GPAS is freely accessible to all academic users at https://gpas.nh.ac.cn. Curated metadata for all projects and the analysis codes are available on GitHub at https://github.com/GPAS-Team. All data produced in the present study are available upon reasonable requests to the authors.

## Authors’ contributions

X. Zhang, Tao. Li, T. Zhou, A. Li, and T. T. Li conceptualized and supervised the study. T. T. Li, H. Hong, and D. Fan performed the methodological study, including algorithm development, data analysis, and AI model training. Tao. Li, T. T. Li, H. Hong, D. Fan, J. Li, J. Wu, S. Jiang, H. Yuan, W. Zheng, and H. Ma developed the data analysis system. Ting. Li, Z. Liu, Z. Su, X. Yu, Y. Liu, X. Li, Y. Shen, C. Zhang, Y. Wang, M. Ma, K. Zhang, F. Du, and Y. Yi obtained clinical samples and performed NGS sequencing and data processing. J. Li, Ting. Li, Z. Liu, Z. Su, and X. Yu conducted functional testing and data analysis of the GPAS system and algorithm. J. Li, Ting. Li, X. Xie, Yawei. Zhang, M. Hu, X. Yin, Yizhe. Zhang, B. Zhao, and H. Liu conducted real-world big data analysis testing of the GPAS system and algorithm. Ting. Li, J. Man, L. Sun, Q. Han, J. Chen, L. Chen, N. Wang, W. Li, K. He, and F. Dong designed the experimental validation strategies and obtained experimental data for algorithm validation and GPAS system function testing. X. Zhang, T. T. Li, Tao. Li, T. Zhou, A. Li, H. Hong, D. Fan, and Y. Yi acquired fundings for the study and wrote the manuscript.

## Acknowledgements

This work was supported by State Key Laboratory of Biomedical Analysis and grants from the China National Natural Science Foundation (No. 82550131, No. 81925017 and No. 82130052 to Tao Li, No. 62503496 to Hao Hong). We would like to thank Professor Xiuwu Bian, Ruifu Yang, Tingbo Liang, Wei Liu, Boan Li, Junchang Cui and Xiaoai Zhang for their help in collecting samples and valuable suggestions.

## Reference

1 Torres Montaguth, O. E., Buddle, S., Morfopoulou, S. & Breuer, J. Clinical metagenomics for diagnosis and surveillance of viral pathogens. Nat Rev Microbiol 24, 61–75 (2026). 10.1038/s41579-025-01223-5

2 Zhou, T. & Zhao, F. AI-empowered human microbiome research. Gut (2025). 10.1136/gutjnl-2025-335946

3 Tegegne, H. A. & Savidge, T. C. Gut microbiome metagenomics in clinical practice: bridging the gap between research and precision medicine. Gut Microbes 17, 2569739 (2025). 10.1080/19490976.2025.2569739

4 Liu, S. et al. Analysis of metagenomic data. Nat Rev Methods Primers 5 (2025). 10.1038/s43586-024-00376-6

5 Ko, K. K. K., Chng, K. R. & Nagarajan, N. Metagenomics-enabled microbial surveillance. Nat Microbiol 7, 486–496 (2022). 10.1038/s41564-022-01089-w

6 Sun, Z. et al. Challenges in benchmarking metagenomic profilers. Nat Methods 18, 618–626 (2021). 10.1038/s41592-021-01141-3

7 Ye, S. H., Siddle, K. J., Park, D. J. & Sabeti, P. C. Benchmarking Metagenomics Tools for Taxonomic Classification. Cell 178, 779–794 (2019). 10.1016/j.cell.2019.07.010

8 Breitwieser, F. P., Lu, J. & Salzberg, S. L. A review of methods and databases for metagenomic classification and assembly. Brief Bioinform 20, 1125–1136 (2019). 10.1093/bib/bbx120

9 Wood, D. E., Lu, J. & Langmead, B. Improved metagenomic analysis with Kraken 2. Genome Biol 20, 257 (2019). 10.1186/s13059-019-1891-0

10 Jenike, K. M. et al. k-mer approaches for biodiversity genomics. Genome Res 35, 219–230 (2025). 10.1101/gr.279452.124

11 Marti, J. M. et al. Addressing the dynamic nature of reference data: a new nucleotide database for robust metagenomic classification. mSystems 10, e0123924 (2025). 10.1128/msystems.01239-24

12 Shaw, J. & Yu, Y. W. Rapid species-level metagenome profiling and containment estimation with sylph. Nat Biotechnol (2024). 10.1038/s41587-024-02412-y

13 Ondov, B. D. et al. Mash: fast genome and metagenome distance estimation using MinHash. Genome Biol 17, 132 (2016). 10.1186/s13059-016-0997-x

14 Pierce, N. T., Irber, L., Reiter, T., Brooks, P. & Brown, C. T. Large-scale sequence comparisons with sourmash. F1000Res 8, 1006 (2019). 10.12688/f1000research.19675.1

15 Acharya, D. B., Kuppan, K. & Divya, B. Agentic AI: Autonomous intelligence for complex goals—A comprehensive survey. IEEe Access 13, 18912–18936 (2025).

16 Wang, C. et al. A survey for large language models in biomedicine. Artif Intell Med 170, 103268 (2025). 10.1016/j.artmed.2025.103268

17 Nguyen, E. et al. Sequence modeling and design from molecular to genome scale with Evo. Science 386, eado9336 (2024). 10.1126/science.ado9336

18 Hou, X. et al. Using artificial intelligence to document the hidden RNA virosphere. Cell 187, 6929–6942 e6916 (2024). 10.1016/j.cell.2024.09.027

19 Ji, Y., Zhou, Z., Liu, H. & Davuluri, R. V. DNABERT: pre-trained Bidirectional Encoder Representations from Transformers model for DNA-language in genome. Bioinformatics 37, 2112–2120 (2021). 10.1093/bioinformatics/btab083

20 Karkera, N., Acharya, S. & Palaniappan, S. K. Leveraging pre-trained language models for mining microbiome-disease relationships. BMC Bioinformatics 24, 290 (2023). 10.1186/s12859-023-05411-z

21 Jiang, P. et al. Adaptation of agentic ai. arXiv preprint arXiv:2512.16301 (2025).

22 Tu, T. et al. Towards conversational diagnostic artificial intelligence. Nature 642, 442–450 (2025).

23 Chorlton, S. D. Ten common issues with reference sequence databases and how to mitigate them. Front Bioinform 4, 1278228 (2024). 10.3389/fbinf.2024.1278228

24 Jain, C., Rodriguez, R. L., Phillippy, A. M., Konstantinidis, K. T. & Aluru, S. High throughput ANI analysis of 90K prokaryotic genomes reveals clear species boundaries. Nat Commun 9, 5114 (2018). 10.1038/s41467-018-07641-9

25 Li, H. Minimap2: pairwise alignment for nucleotide sequences. Bioinformatics 34, 3094–3100 (2018). 10.1093/bioinformatics/bty191

26 Song, L. & Langmead, B. Centrifuger: lossless compression of microbial genomes for efficient and accurate metagenomic sequence classification. Genome Biol 25, 106 (2024). 10.1186/s13059-024-03244-4

27 Piro, V. C. & Reinert, K. ganon2: up-to-date and scalable metagenomics analysis. (2024). 10.1101/2023.12.07.570547

28 Meyer, F. et al. Critical Assessment of Metagenome Interpretation: the second round of challenges. Nat Methods 19, 429–440 (2022). 10.1038/s41592-022-01431-4

29 Liu, Y., Elworth, R. A. L., Jochum, M. D., Aagaard, K. M. & Treangen, T. J. De novo identification of microbial contaminants in low microbial biomass microbiomes with Squeegee. Nat Commun 13, 6799 (2022). 10.1038/s41467-022-34409-z

30 Guo, D. et al. DeepSeek-R1 incentivizes reasoning in LLMs through reinforcement learning. Nature 645, 633–638 (2025). 10.1038/s41586-025-09422-z

31 Wu, G. et al. A core microbiome signature as an indicator of health. Cell 187, 6550–6565 e6511 (2024). 10.1016/j.cell.2024.09.019

32 van Vollenhoven, R. F. et al. A Phase 3 Trial of Telitacicept for Systemic Lupus Erythematosus. N Engl J Med 393, 1475–1485 (2025). 10.1056/NEJMoa2414719

33 Fourgeaud, J. et al. Performance of clinical metagenomics in France: a prospective observational study. Lancet Microbe 5, e52–e61 (2024). 10.1016/S2666-5247(23)00244-6

34 Zhang, Z. et al. Assessment of global health risk of antibiotic resistance genes. Nat Commun 13, 1553 (2022). 10.1038/s41467-022-29283-8

## Reference

35 Li, H. et al. The Sequence Alignment/Map format and SAMtools. Bioinformatics 25, 2078–2079 (2009). 10.1093/bioinformatics/btp352

36 Shaw, J. & Yu, Y. W. Fast and robust metagenomic sequence comparison through sparse chaining with skani. Nat Methods 20, 1661–1665 (2023). 10.1038/s41592-023-02018-3

37 Zielezinski, A. et al. Ultrafast and accurate sequence alignment and clustering of viral genomes. Nat Methods 22, 1191–1194 (2025). 10.1038/s41592-025-02701-7

38 Morgulis, A., Gertz, E. M., Schaffer, A. A. & Agarwala, R. A fast and symmetric DUST implementation to mask low-complexity DNA sequences. J Comput Biol 13, 1028–1040 (2006). 10.1089/cmb.2006.13.1028

39 Henriksen, R. A., Zhao, L. & Korneliussen, T. S. NGSNGS: next-generation simulator for next-generation sequencing data. Bioinformatics 39 (2023). 10.1093/bioinformatics/btad041

40 Ye, G. et al. MAGdb: a comprehensive high quality MAGs repository for exploring microbial metagenome-assemble genomes. Genome Biol 26, 276 (2025). 10.1186/s13059-025-03711-6

