## Supplemental Figures for "GPAS: an online AI system for rapid and accurate pathogen identification and LLM-based interpretation"

**a**

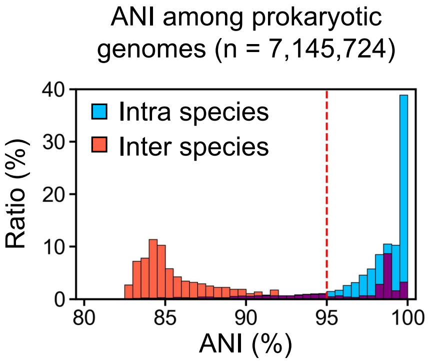


**b**

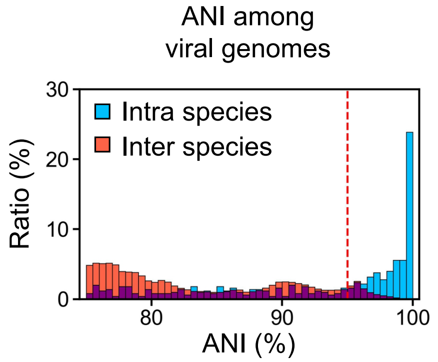


**c**

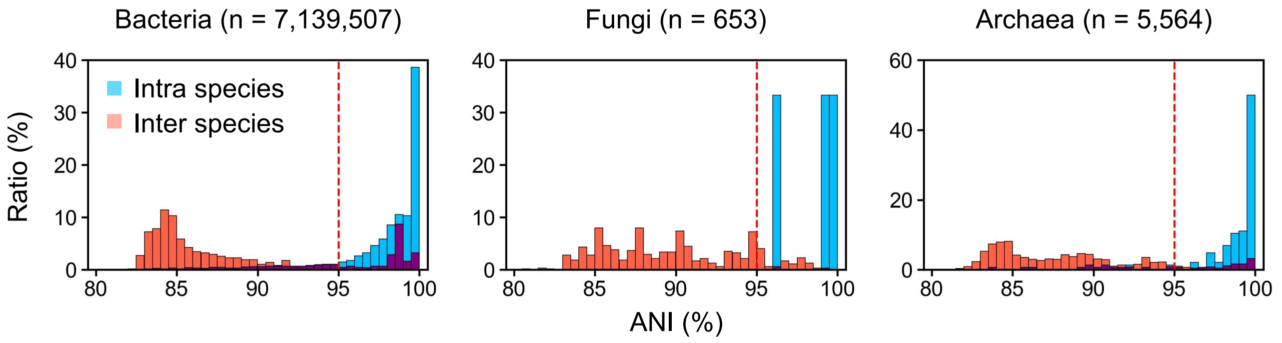


**d**

Inter-species genome pairs with high ANI

| Domain | Inter-species pairs (> 95% ANI) | Inter-species pairs (> 99% ANI) |
|---|---|---|
| bacteria | 1,417,553 | 349,652 |
| fungi | 79 | 2 |
| archaea | 650 | 261 |
| virus | 21,046 | 430 |

**e**

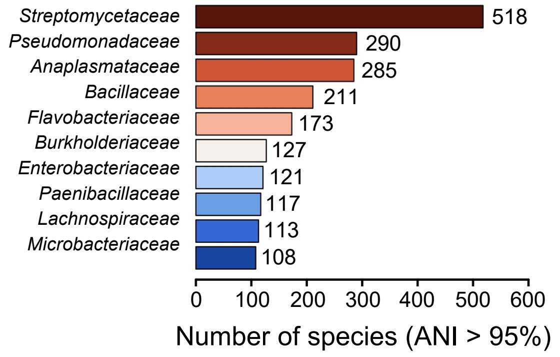


**f**

ANI of Enterobacteriaceae family (845 species, taxonomy ID: 543)

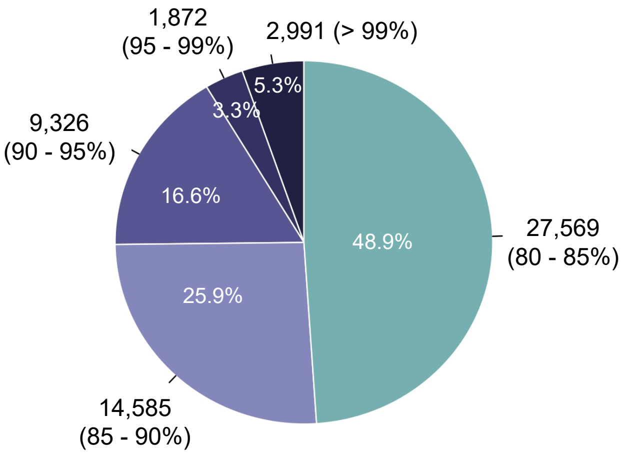


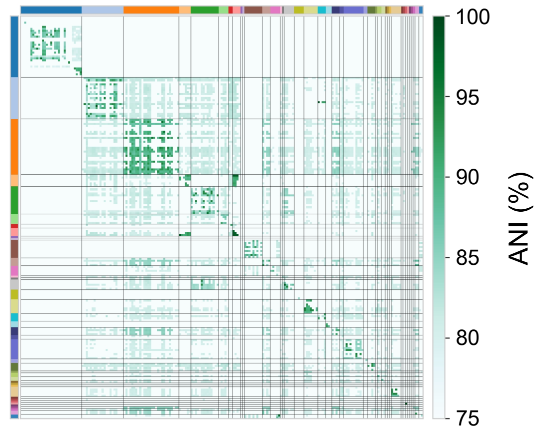

**Extended Data Fig. 1 Genome similarity analysis across microbes.** a-d, Genome-wide sequence similarity analysis across different microbial taxa. Bar plots display average nucleotide identity (ANI) distributions within and between species for bacteria, archaea, fungi, and viruses, revealing extensive genomic homology particularly among closely related species. e. Top 10 families that contain the most high-ANI species. f. ANI distribution of Enterobacteriaceae family.

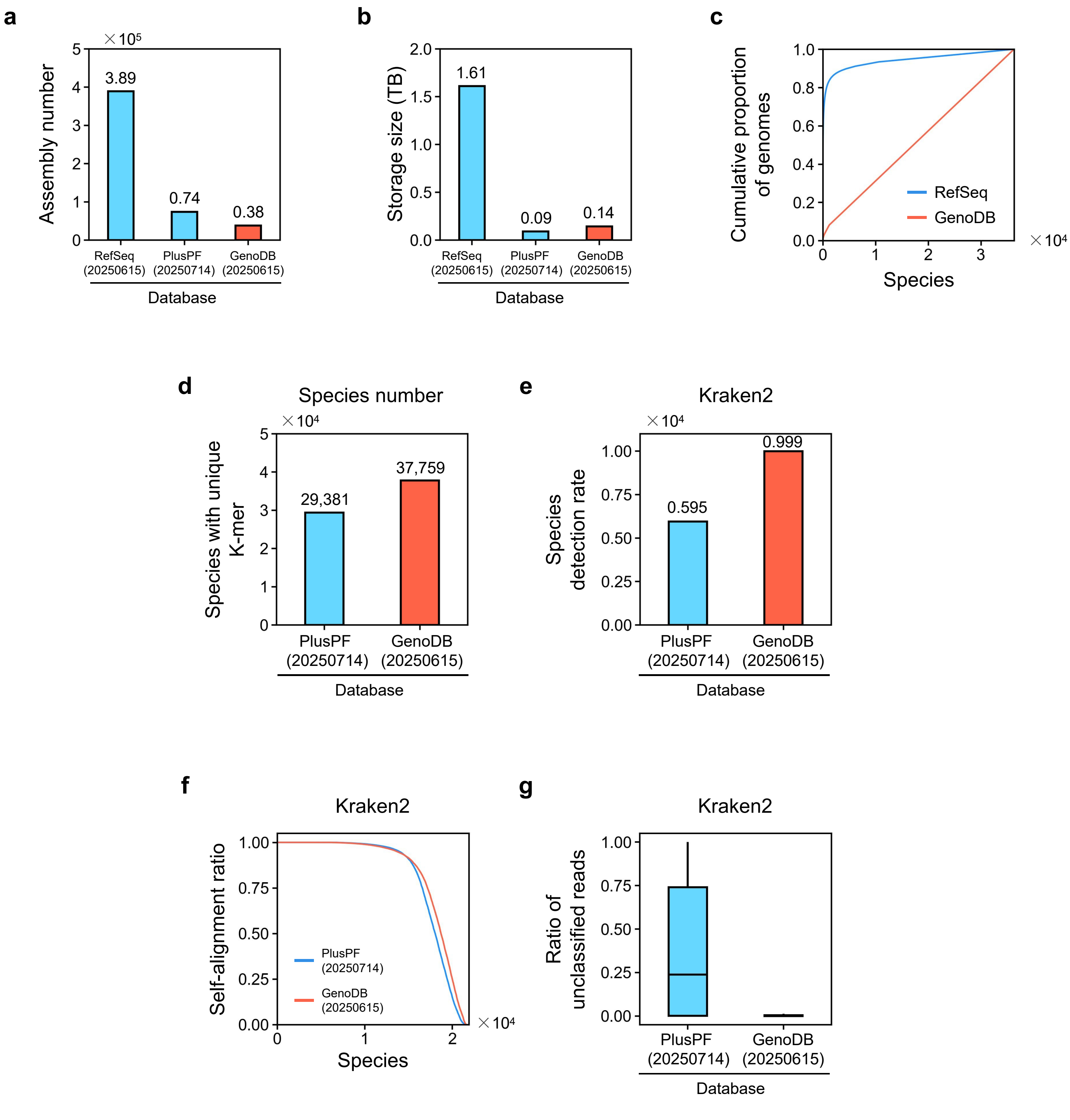


**Extended Data Fig. 2 Construction of the non-redundant microbial database GenoDB.** a-c, Demonstration of database compression efficiency. GenoDB reduces the overall database size to one-tenth of the original volume through representative genome selection. d-g, Performance comparison between GenoDB and the Kraken2 default PlusPF database. GenoDB achieves broader species coverage, higher self-mapping rates, and lower unclassified read proportions across simulated datasets, demonstrating a favorable balance between data completeness and computational efficiency.

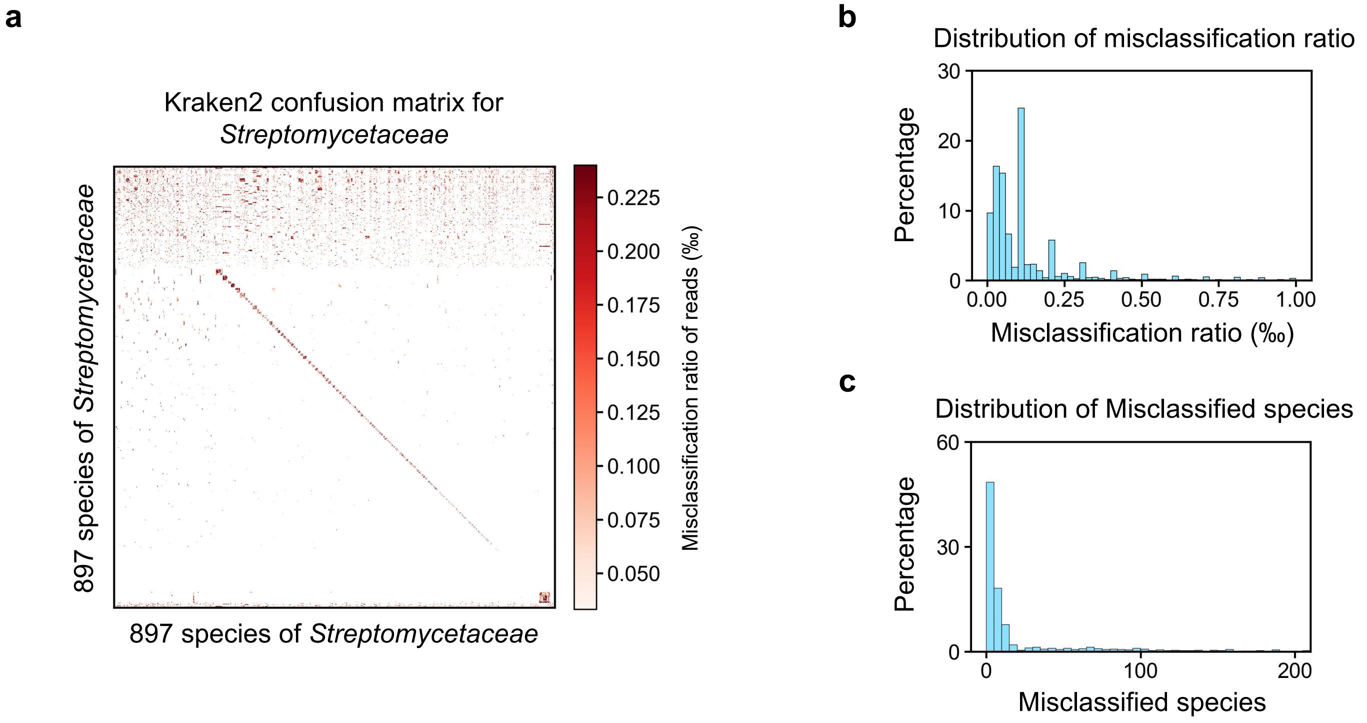


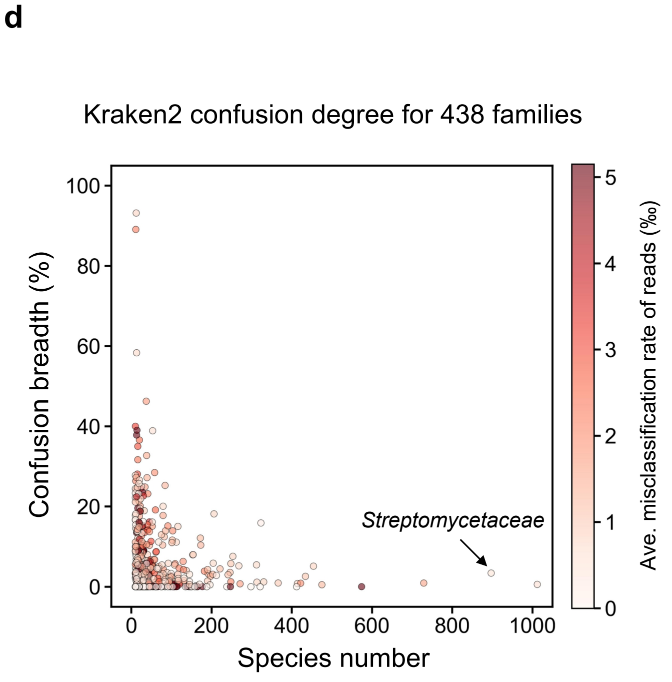


**e**

Top 10 families with highest confusion degree in Kraken2

| Family | Sp. number | Confusion breadth (%) | Ave. confusion ratio (‰) |
|---|---|---|---|
| *Pectobacteriaceae* | 58 | 28.5 | 2.0 |
| *Aeromonadaceae* | 60 | 19.2 | 2.7 |
| *Fusobacteriaceae* | 28 | 23.5 | 6.6 |
| *Borreliaceae* | 37 | 46.2 | 1.8 |
| *Bacteroidaceae* | 84 | 25.3 | 1.4 |
| *Brucellaceae* | 45 | 16.2 | 4.0 |
| *Dietziaceae* | 13 | 37.8 | 11.1 |
| *Listeriaceae* | 30 | 22.6 | 3.6 |
| *Thalassospiraceae* | 14 | 39.0 | 7.1 |
| *Coriobacteriaceae* | 22 | 18.8 | 8.0 |

**Extended Data Fig. 3 Whole microbiome-wide interspecies misclassification probability matrix calculation.** a, Representative Kraken2 confusion matrix for Streptomycetaceae (897 species), showing the misclassification ratio of reads (‰) across species pairs. b-c, Distribution of misclassification ratios and misclassified species counts across streptomycetaceae family. d, Confusion breadth percentage for 438 bacterial families, illustrating the extent of cross-species read misassignment. e, Top 10 families with the highest confusion degree in Kraken2, listing family name, species number, confusion breadth (%), and average confusion ratio (‰).

**a**

Kraken2 inter-species misclassification categories

| Species Categories | Sub-categories & annotation | Species number (10x simu-reads) | Species number (1x simu-reads) | Species number (0.1x simu-reads) |
|---|---|---|---|---|
| **Primarily self-reported** | **Exclusive Self-Assignment:** All simulated reads were correctly assigned only to the species of origin, with no cross-species misassignment to it. | 11,108 | 14,597 | 20,391 |
| **Cross-reported** | **Predominant Self-Assignment:** Reads were assigned to multiple species, but the highest proportion was to the correct species. | 16,362 | 12,734 | 7,046 |
| | **Non-Predominant Self-Assignment:** Reads were assigned to multiple species, with the correct species not receiving the highest proportion. | 1,921 | 1,319 | 885 |
| | **Low-Rate Self-Assignment (<10%):** Less than 10% of reads were correctly assigned, with most assigned to other species. | 748 | 461 | 266 |
| **Non-self-reported** | **No Assignment:** All reads failed to be assigned to any species.<br>**Complete Misassignment:** All reads were assigned to other species, with none assigned to the correct species. | 386 | 465 | 805 |

**b**

Sylph inter-species misclassification categories

| Assembly categories | Assembly number (10x simu-reads) | Assembly number (1x simu-reads) | Assembly number (0.1x simu-reads) |
|---|---|---|---|
| **Match Same Assembly Only** | 49,176 | 42,700 | 27,127 |
| **Match Another Same Species Assembly Only** | 2,430 | 7,810 | 15,737 |
| **Match Another Different Species Assembly Only** | 59 | 289 | 790 |
| **Match Same Assembly And Beside** | 823 | 974 | 515 |
| **Match Another Same Species Assembly And Beside** | 80 | 773 | 607 |
| **Match Different Species Assembly** | 0 | 2 | 16 |
| **Match NULL** | 5,530 | 5,531 | 12,716 |

**Extended Data Fig. 4 Detailed inter-species misclassification categories for Kraken2 and Sylph.** a, Kraken2 inter-species misclassification categories across three sequencing depths (10×, 1×, 0.1×). b, Sylph inter-species misclassification categories at the assembly level across three sequencing depths.

**a**

The composition of 40,000 samples used for training the GPAS model

| Dataset Name | Mixed Containments | | | Sample Number |
|---|---|---|---|---|
| | Coverages (x) | Domains | Species Number | |
| All_01 | 0.1, 0.01 | Archaea, Bacteria, Fungi, Viral | 2 | 1000 |
| All_02 | 10, 0.01 | Archaea, Bacteria, Fungi, Viral | 2 | 1000 |
| All_03 | 1, 0.01 | Archaea, Bacteria, Fungi, Viral | 2 | 1000 |
| All_04 | 3, 0.01 | Archaea, Bacteria, Fungi, Viral | 2 | 1000 |
| All_05 | 10, 0.1, 0.01 | Archaea, Bacteria, Fungi, Viral | 3 | 1000 |
| All_06 | 10, 1, 0.01 | Archaea, Bacteria, Fungi, Viral | 3 | 1000 |
| All_07 | 10, 3, 0.01 | Archaea, Bacteria, Fungi, Viral | 3 | 1000 |
| All_08 | 1, 0.1, 0.01 | Archaea, Bacteria, Fungi, Viral | 3 | 1000 |
| All_09 | 10, 3, 1, 0.1, 0.01 | Archaea, Bacteria, Fungi, Viral | 5 | 5000 |
| All_10 | 10, 1, 0.1, 0.01 | Archaea, Bacteria, Fungi, Viral | 10 | 1000 |
| All_11 | 10, 3, 0.1, 0.01 | Archaea, Bacteria, Fungi, Viral | 10 | 1000 |
| All_12 | 10, 3, 1, 0.01 | Archaea, Bacteria, Fungi, Viral | 10 | 1000 |
| All_13 | 10, 3, 1, 0.1, 0.01 | Archaea, Bacteria, Fungi, Viral | 10 | 5000 |
| Bac_01 | 10, 3 | Bacteria | 2 | 1000 |
| Bac_02 | 10, 1 | Bacteria | 2 | 1000 |
| Bac_03 | 10, 0.1 | Bacteria | 2 | 1000 |
| Bac_04 | 10, 3, 1 | Bacteria | 3 | 1000 |
| Bac_05 | 10, 3, 0.1 | Bacteria | 3 | 1000 |
| Bac_06 | 10, 1, 0.1 | Bacteria | 3 | 1000 |
| Bac_07 | 10, 3, 1, 0.1 | Bacteria | 10 | 1000 |
| Viral_01 | 10, 3, 1 | Viral | 3 | 3000 |
| Viral_02 | 10, 3, 1, 0.1 | Viral | 4 | 3000 |
| Viral_03 | 10, 3, 1, 0.1, 0.01 | Viral | 5 | 3000 |
| Viral_04 | 10, 3, 1, 0.1, 0.01 | Viral | 10 | 3000 |

**b**

Training performance of the GPAS model

| | | Precision | Recall | F1 | TP | FP | FN |
|---|---|---|---|---|---|---|---|
| Assembly Level | Sylph | 0.831 | **0.493** | 0.619 | 106,595 | 21,699 | 109,405 |
| | GPAS Anchored | **0.944** | 0.484 | **0.640** | 104,558 | 6,233 | 111,442 |
| Species Level | Sylph | 0.972 | **0.579** | **0.726** | 123,885 | 3,583 | 90,125 |
| | GPAS Anchored | **0.995** | 0.514 | 0.677 | 109,905 | 546 | 104,105 |

**Extended Data Fig. 5 All 40,000 samples used for training the GPAS model.** a, Composition of the 40,000 simulated metagenomic samples used for model training and validation. The dataset encompasses diverse combinations of microbial domains (archaea, bacteria, fungi, viruses), species numbers, and sequencing depths (ranging from 0.01× to 10×), with sample counts specified for each configuration. b, Training performance of the GPAS model at both assembly and species levels. Precision, recall, F1-score, true positives (TP), false positives (FP), and false negatives (FN) are reported for Sylph baseline and GPAS anchored results, demonstrating substantial improvement in precision with GPAS calibration.

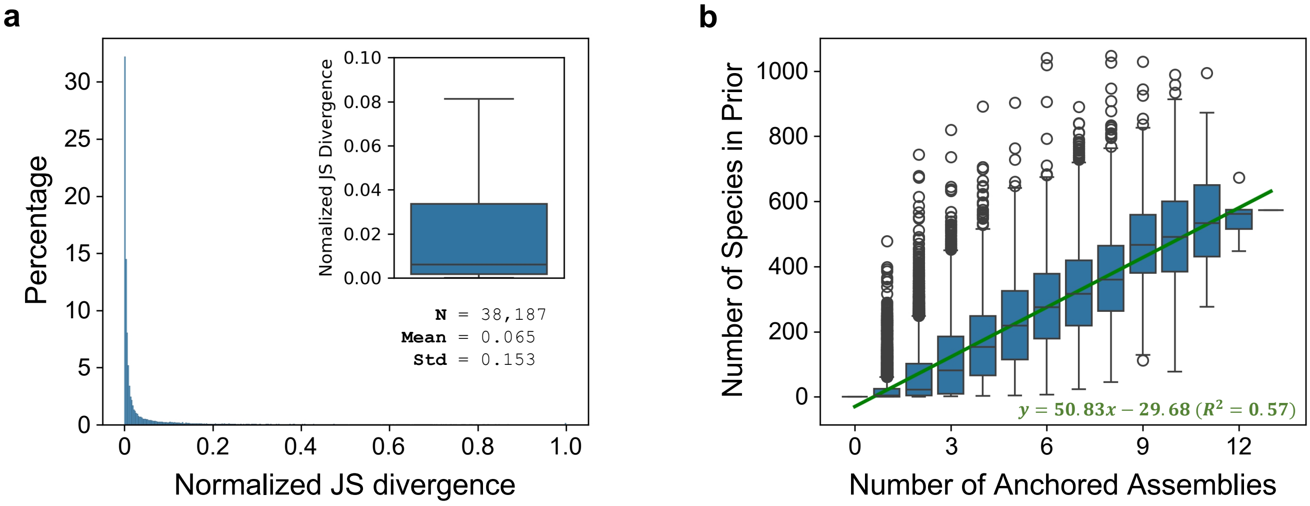


**Extended Data Fig. 6 Anchor-based mixture prior building performance versus ground truth observation.** a, Comparison of the mixture prior abundance distribution constructed from anchor species against the ground truth observation. The normalized Jensen-Shannon divergence (mean 0.065, variance 0.153) indicates excellent agreement between predicted and observed abundance distributions. b, Average number of false-positive species eliminated per anchor species (mean 50), demonstrating the effectiveness of anchor-based calibration in reducing false identifications.

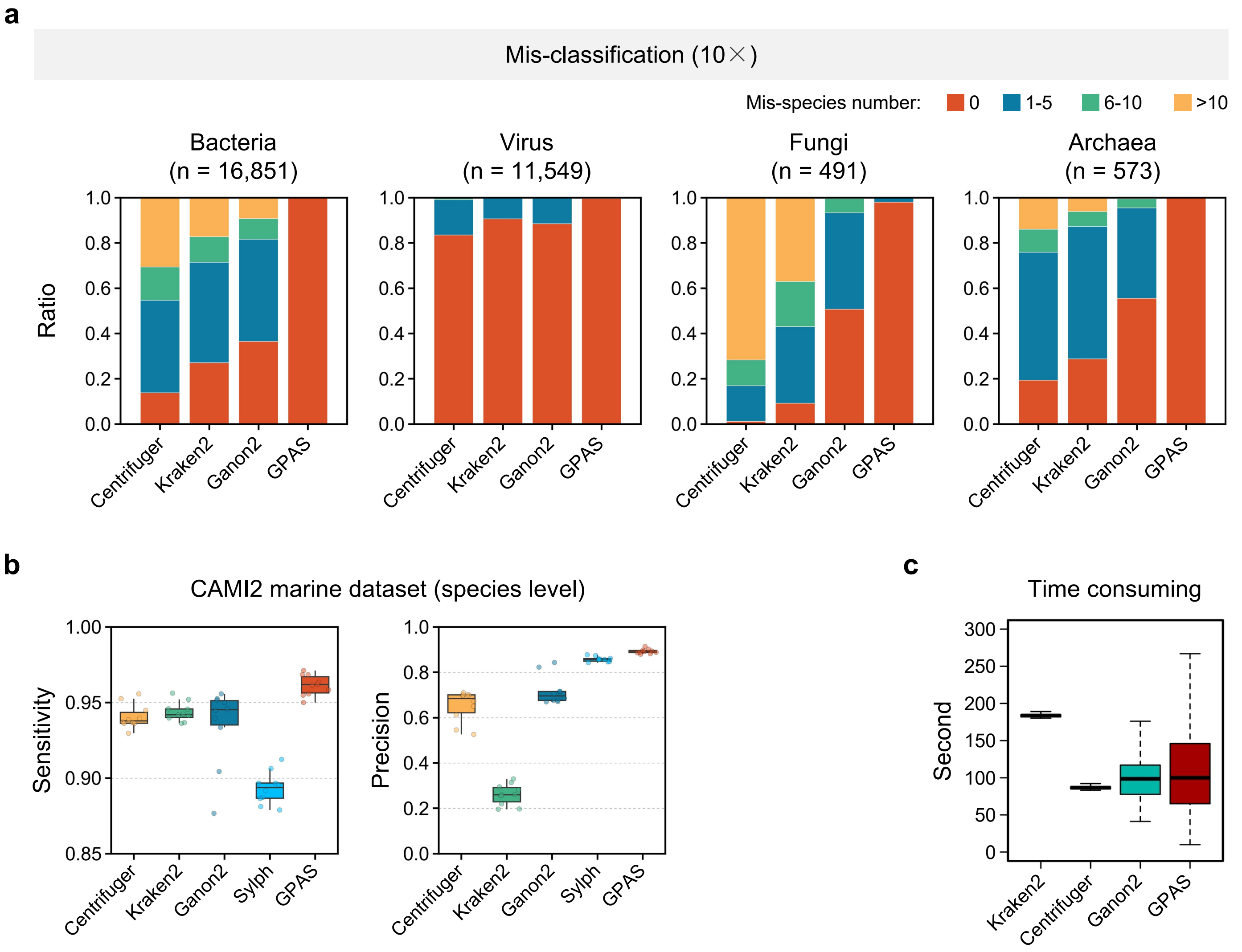


**Extended Data Fig. 7 Performance of GPAS compared to other microbe profiling tools.** a, Detailed comparison of false-positive rates at 10× sequencing depths for GPAS and mainstream tools (Centrifuger, Kraken2, Ganon2). GPAS consistently achieves the lowest false-positive counts across all depths. b, Comprehensive performance metrics (precision, recall) on the CAMI II marine metagenome dataset, showing GPAS's superior performance compared to Centrifuger, Kraken2, and Ganon2. c, Runtime comparison across tools. By preloading the GenoDB database into memory, GPAS achieves computational efficiency.

**a**

| Category | Sample number | Description |
|---|---|---|
| Negative control | 100 | Genomic DNA of A549 cell line |
| Positive control | 100 | Mixture of genomic DNA of *Staphylococcus aureus* and A549 |
| blood | 100 | NGS raw data of 1000g project |

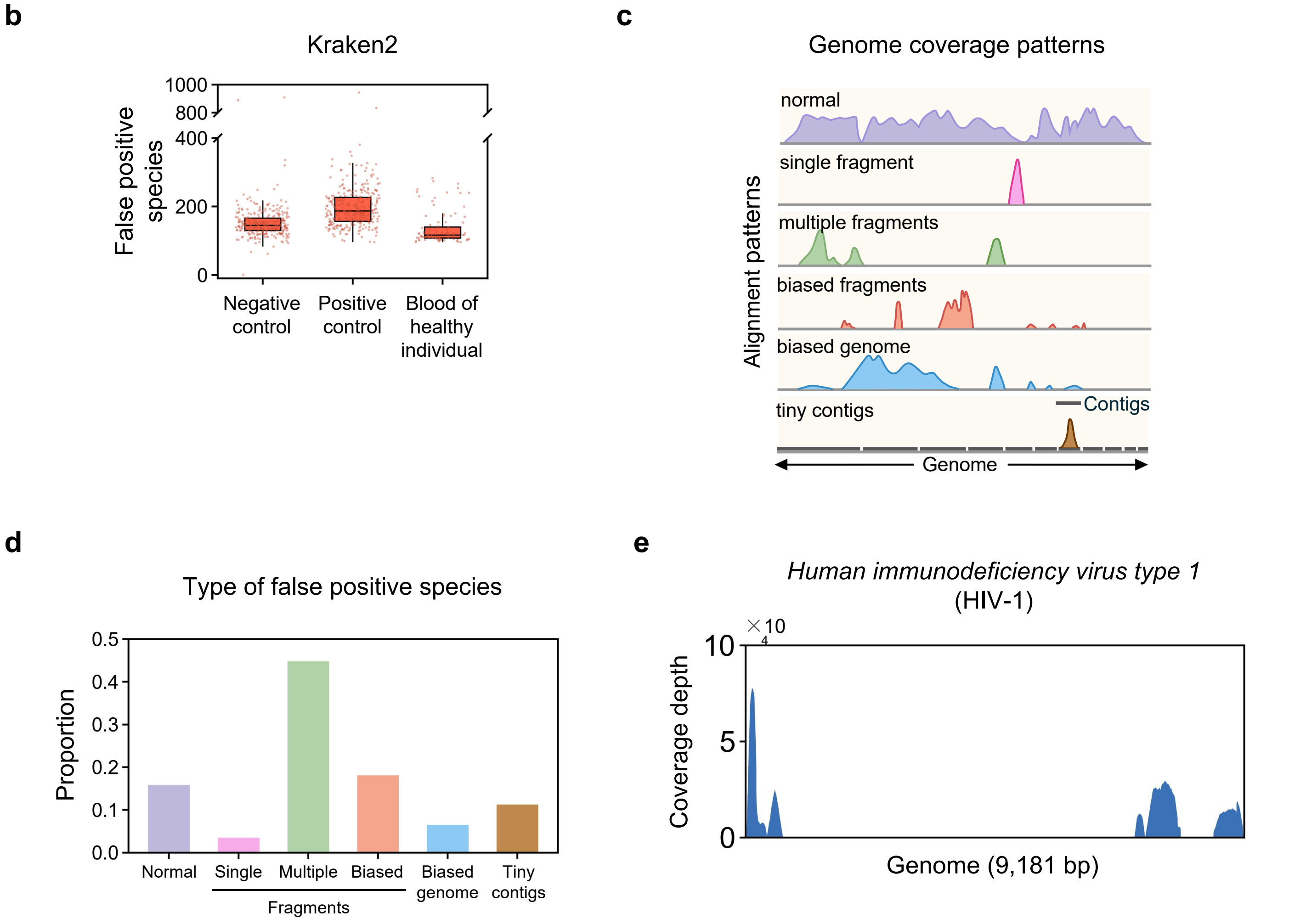


**Extended Data Fig. 8 Genome coverage characteristics of microbial species.** a, Sample categories used for coverage pattern analysis, including negative controls (A549 cell line genomic DNA, n=100), positive controls (mixture of Staphylococcus aureus and A549 genomic DNA, n=100), and blood samples from the 1000 Genomes Project (n=100). b-d, Classification of false-positive species types and their characteristic genome coverage patterns. e, Representative genome coverage profile of Human immunodeficiency virus type 1 (HIV-1) from a real metagenomic sample, illustrating the fragmented, biased distribution typical of false-positive identifications.

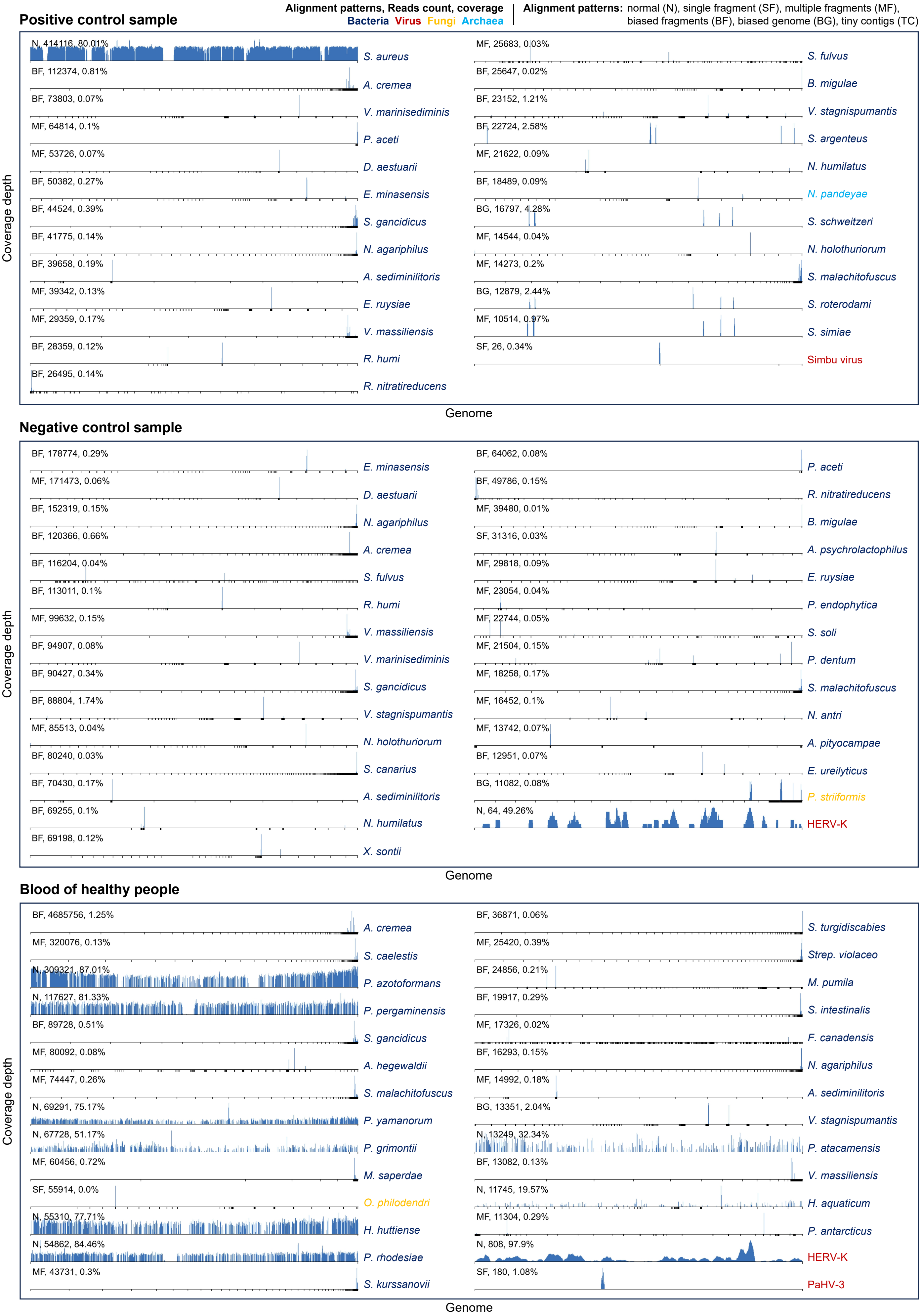
Alignment patterns, Reads count, coverage
Bacteria Virus Fungi Archaea
Alignment patterns: normal (N), single fragment (SF), multiple fragments (MF), biased fragments (BF), biased genome (BG), tiny contigs (TC)
Positive control sample
N, 414116, 80.01% S. aureus
BF, 112374, 0.81% A. cremea
BF, 73803, 0.07% V. marinisediminis
MF, 64814, 0.1% P. aceti
MF, 53726, 0.07% D. aestuarii
BF, 50382, 0.27% E. minasensis
BF, 44524, 0.39% S. gancidicus
BF, 41775, 0.14% N. agariphilus
BF, 39658, 0.19% A. sediminilitoris
MF, 39342, 0.13% E. ruysiae
MF, 29359, 0.17% V. massiliensis
BF, 28359, 0.12% R. humi
BF, 26495, 0.14% R. nitratireducens
MF, 25683, 0.03% S. fulvus
BF, 25647, 0.02% B. migulae
BF, 23152, 1.21% V. stagnispumantis
BF, 22724, 2.58% S. argenteus
MF, 21622, 0.09% N. humilatus
BF, 18489, 0.09% N. pandeyae
BG, 16797, 4.28% S. schweitzeri
MF, 14544, 0.04% N. holothuriorum
MF, 14273, 0.2% S. malachitofuscus
BG, 12879, 2.44% S. roterodami
MF, 10514, 0.97% S. simiae
SF, 26, 0.34% Simbu virus
Coverage depth
Genome
Negative control sample
BF, 178774, 0.29% E. minasensis
MF, 171473, 0.06% D. aestuarii
BF, 152319, 0.15% N. agariphilus
BF, 120366, 0.66% A. cremea
BF, 116204, 0.04% S. fulvus
BF, 113011, 0.1% R. humi
MF, 99632, 0.15% V. massiliensis
BF, 94907, 0.08% V. marinisediminis
BF, 90427, 0.34% S. gancidicus
BF, 88804, 1.74% V. stagnispumantis
MF, 85513, 0.04% N. holothuriorum
BF, 80240, 0.03% S. canarius
BF, 70430, 0.17% A. sediminilitoris
BF, 69255, 0.1% N. humilatus
BF, 69198, 0.12% X. sontii
BF, 64062, 0.08% P. aceti
BF, 49786, 0.15% R. nitratireducens
MF, 39480, 0.01% B. migulae
SF, 31316, 0.03% A. psychrolactophilus
MF, 29818, 0.09% E. ruysiae
MF, 23054, 0.04% P. endophytica
MF, 22744, 0.05% S. soli
MF, 21504, 0.15% P. dentum
MF, 18258, 0.17% S. malachitofuscus
MF, 16452, 0.1% N. antri
MF, 13742, 0.07% A. pityocampae
BF, 12951, 0.07% E. ureilyticus
BG, 11082, 0.08% P. striiformis
N, 64, 49.26% HERV-K
Coverage depth
Genome
Blood of healthy people
BF, 4685756, 1.25% A. cremea
MF, 320076, 0.13% S. caelestis
N, 309321, 87.01% P. azotoformans
N, 117627, 81.33% P. pergaminensis
BF, 89728, 0.51% S. gancidicus
MF, 80092, 0.08% A. hegewaldii
MF, 74447, 0.26% S. malachitofuscus
N, 69291, 75.17% P. yamanorum
N, 67728, 51.17% P. grimontii
MF, 60456, 0.72% M. saperdae
SF, 55914, 0.0% O. philodendri
N, 55310, 77.71% H. huttiense
N, 54862, 84.46% P. rhodesiae
MF, 43731, 0.3% S. kurssanovii
BF, 36871, 0.06% S. turgidiscabies
MF, 25420, 0.39% Strep. violaceo
BF, 24856, 0.21% M. pumila
BF, 19917, 0.29% S. intestinalis
MF, 17326, 0.02% F. canadensis
BF, 16293, 0.15% N. agariphilus
MF, 14992, 0.18% A. sediminilitoris
BG, 13351, 2.04% V. stagnispumantis
N, 13249, 32.34% P. atacamensis
BF, 13082, 0.13% V. massiliensis
N, 11745, 19.57% H. aquaticum
MF, 11304, 0.29% P. antarcticus
N, 808, 97.9% HERV-K
SF, 180, 1.08% PaHV-3
Coverage depth
Genome

**Extended Data Fig. 9 Genome coverage profiles in three sample types.** Genome coverage profiles of representative microbial species across three distinct sample types. Each panel displays the coverage depth distribution along the genomic coordinates, revealing sample type-specific patterns and the consistency of true-positive identifications across different biological contexts.

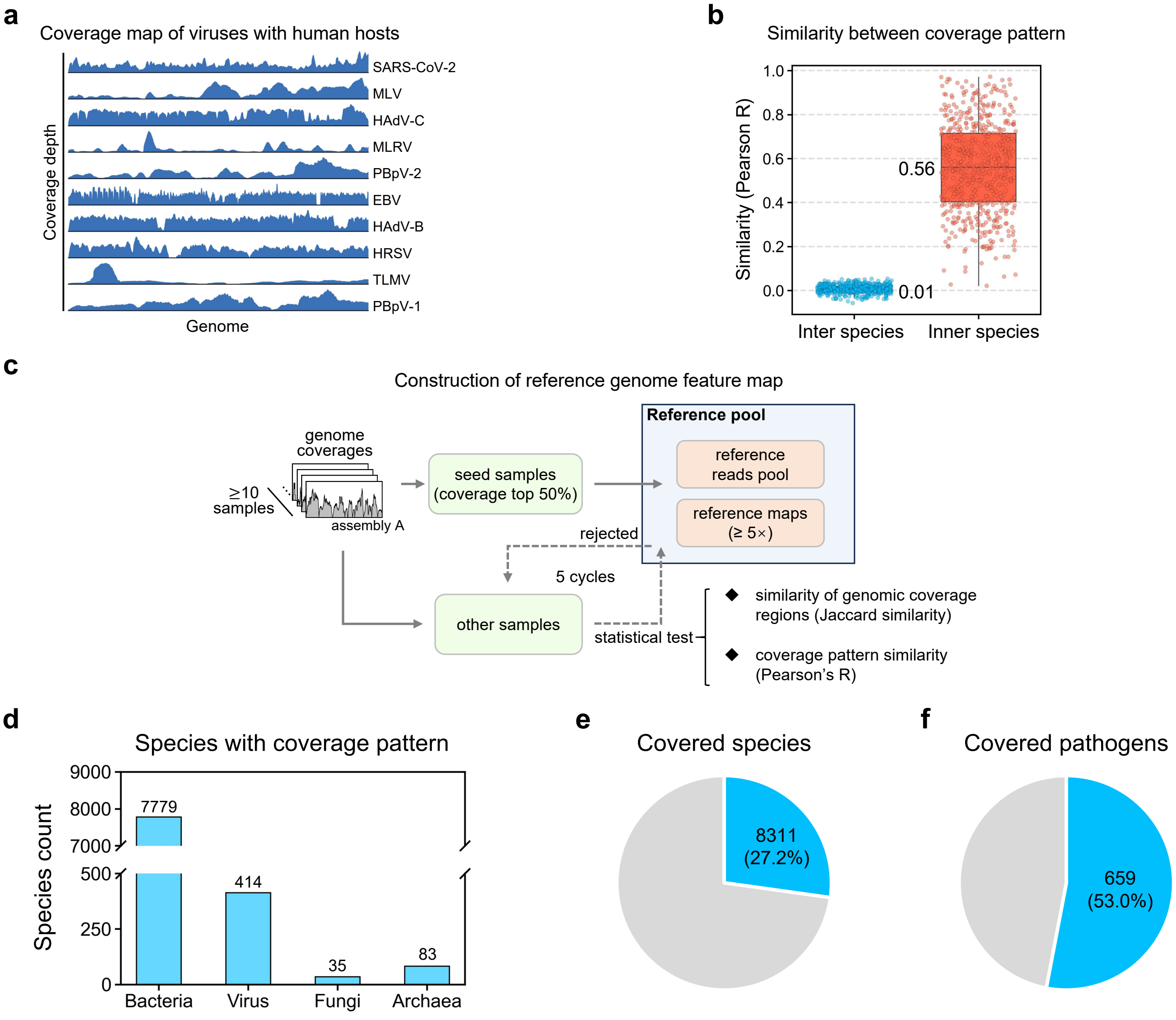


**g**

Data summary for the true positives retained analysis

| GCF_ID | Domain | Name | Detected Samples | Clinically Verified Samples | Selected Samples |
|---|---|---|---|---|---|
| GCF_000733995 | bacteria | Mycoplasmoides pneumoniae | 25 | 2 | 2 |
| GCF_000820495 | virus | Influenza B virus | 38 | 5 | 5 |
| GCF_000848705 | virus | Human respirovirus 1 | 45 | 1 | 1 |
| GCF_000850205 | virus | Human respirovirus 3 | 71 | 5 | 5 |
| GCF_000855545 | virus | Human orthopneumovirus | 78 | 10 | 10 |
| GCF_000865085 | virus | Influenza A virus | 111 | 83 | 37 |
| GCF_000872045 | virus | Human gammaherpesvirus 4 | 509 | 2 | 2 |
| GCF_001343785 | virus | Influenza A virus | 564 | 274 | 188 |
| GCF_001650295 | bacteria | Escherichia coli | 6379 | 1 | 1 |
| GCF_002815475 | virus | Human orthopneumovirus | 32 | 9 | 9 |
| GCF_003425505 | bacteria | Haemophilus influenzae | 981 | 6 | 6 |
| GCF_003522665 | bacteria | Acinetobacter baumannii | 379 | 5 | 5 |
| GCF_003945425 | bacteria | Staphylococcus aureus | 764 | 4 | 4 |
| GCF_009858895 | virus | Severe acute respiratory syndrome coronavirus 2 | 802 | 436 | 267 |
| GCF_013201115 | bacteria | Pseudomonas aeruginosa | 377 | 6 | 6 |
| GCF_017901095 | bacteria | Mycobacterium tuberculosis | 25 | 3 | 3 |
| GCF_018219285 | bacteria | Enterococcus faecium | 911 | 1 | 1 |
| GCF_021117215 | bacteria | Stenotrophomonas maltophilia | 44 | 1 | 1 |
| GCF_022343225 | bacteria | Serratia marcescens | 230 | 1 | 1 |
| GCF_022749215 | bacteria | Klebsiella pneumoniae | 2504 | 6 | 6 |
| GCF_026625945 | bacteria | Streptococcus pneumoniae | 3327 | 9 | 9 |
| **Total** | | | **18,196** | **870** | **569** |

**Extended Data Fig. 10 Characteristics of microbial genome coverage.** a, Coverage maps of viruses with human hosts, displaying genome-wide read depth distributions across multiple clinical samples. b, Analysis shows that genome coverage patterns are highly similar among samples of the same species, but differ substantially between different species. c, Workflow for the construction of the reference genome feature map, reference read pool, and reference coverage maps. d-f, Statistical overview of genome coverage characteristics across species. d, The number of microbial species in which the genome coverage pattern was detected, shown for four major categories. e, The proportion of all species in which the pattern was detected. f, The proportion of pathogenic microorganisms in which the pattern was detected. g, Summary of true positives dataset.

### Extended Data Fig. 11 GPAS big data infrastructure

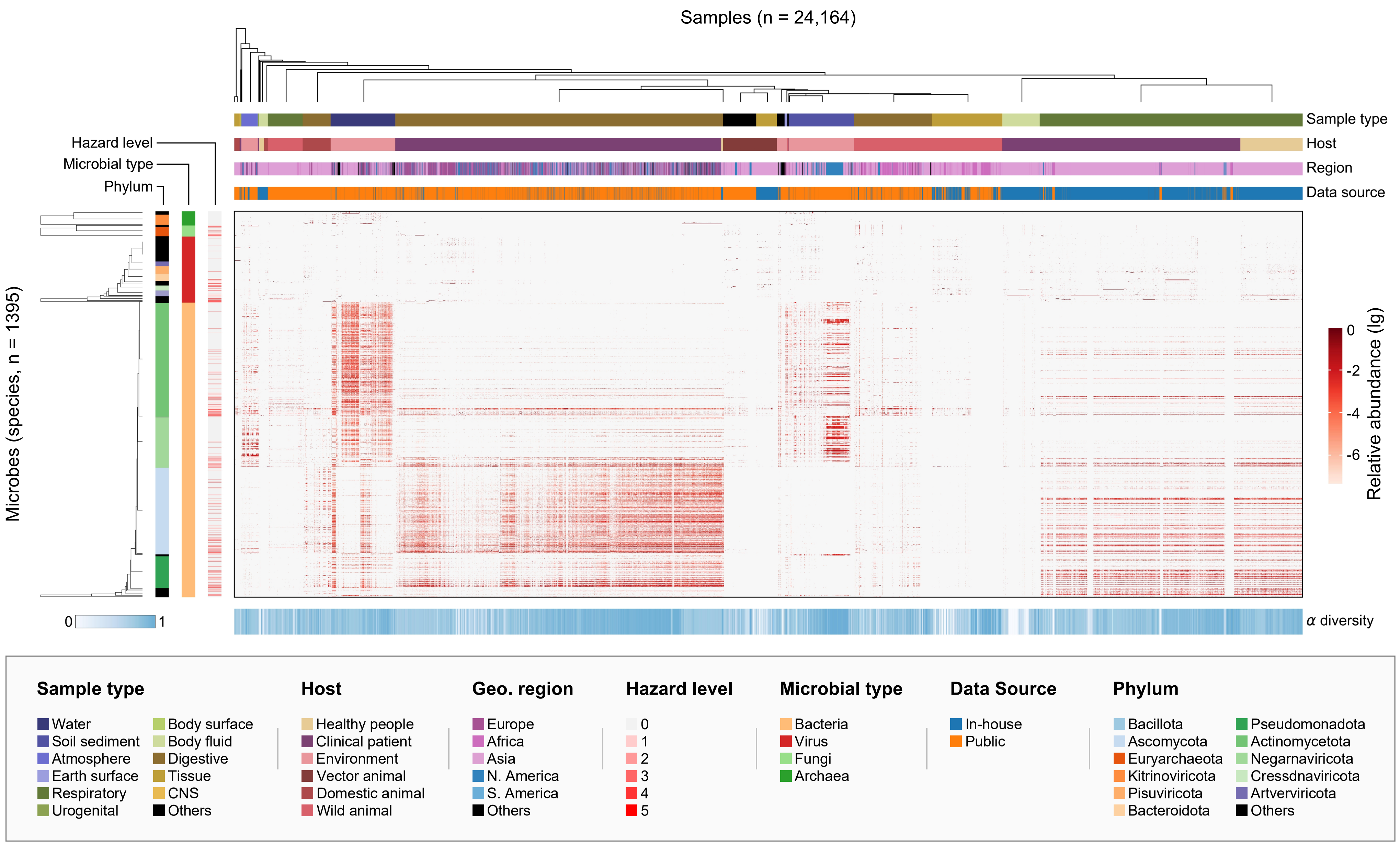

**Extended Data Fig. 11 GPAS big data infrastructure.** Integrated visualization of the 24,164 metagenomic samples constituting GPAS's big data infrastructure. The heatmap displays detected Microbes along with sample metadata including geographic region (S. America, Europe, Africa, Asia, N. America, Others), hazard level (0-5), data source (Public and In-house), microbial type (Bacteria, Virus, Fungi, Archaea), sample type (Body surface, Water, Soil sediment, Atmosphere, Earth surface, Respiratory, Urogenital, Body fluid, Digestive, Tissue, CNS, Others), host category (Healthy people, Vector animal, Domestic animal, Wild animal, Environment, Clinical patient), and phylum-level taxonomy. Relative abundance (log10 scale) is represented by the outer colored bands.

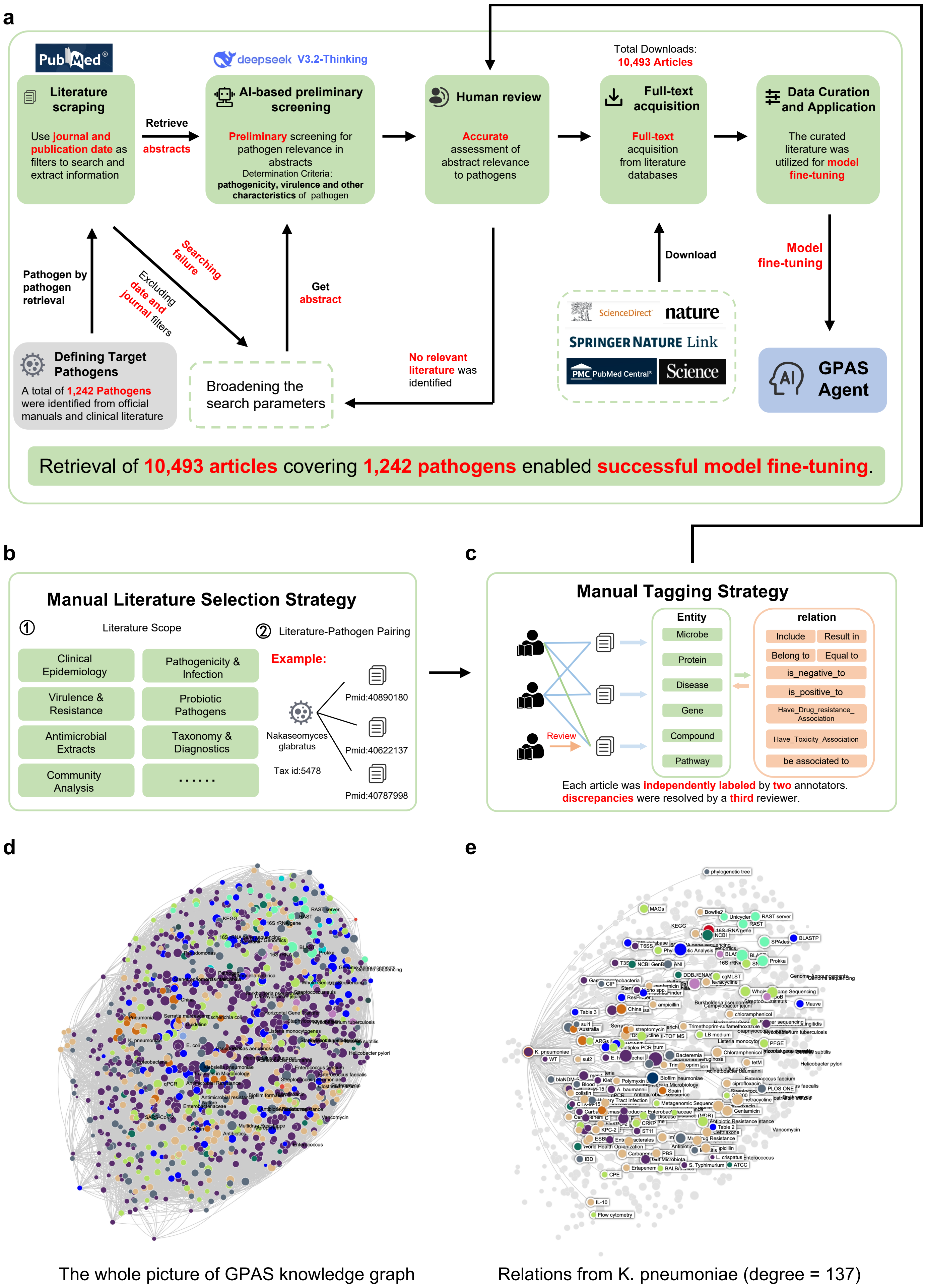
a
PubMed
Literature scraping
Use journal and publication date as filters to search and extract information
Retrieve abstracts
deepseek V3.2-Thinking
AI-based preliminary screening
Preliminary screening for pathogen relevance in abstracts
Determination Criteria: pathogenicity, virulence and other characteristics of pathogen
Human review
Accurate assessment of abstract relevance to pathogens
Total Downloads: 10,493 Articles
Full-text acquisition
Full-text acquisition from literature databases
Data Curation and Application
The curated literature was utilized for model fine-tuning
Pathogen by pathogen retrieval
Searching failure
Excluding date and journal filters
Get abstract
Download
Model fine-tuning
Defining Target Pathogens
A total of 1,242 Pathogens were identified from official manuals and clinical literature
Broadening the search parameters
No relevant literature was identified
ScienceDirect
nature
SPRINGER NATURE Link
PMC PubMed Central®
Science
GPAS Agent
Retrieval of 10,493 articles covering 1,242 pathogens enabled successful model fine-tuning.
b
Manual Literature Selection Strategy
① Literature Scope
② Literature-Pathogen Pairing
Clinical Epidemiology
Pathogenicity & Infection
Virulence & Resistance
Probiotic Pathogens
Antimicrobial Extracts
Taxonomy & Diagnostics
Community Analysis
……
Example:
Nakaseomyces glabratus
Tax id:5478
Pmid:40890180
Pmid:40622137
Pmid:40787998
c
Manual Tagging Strategy
Review
Entity
Microbe
Protein
Disease
Gene
Compound
Pathway
relation
Include
Result in
Belong to
Equal to
is_negative_to
is_positive_to
Have_Drug_resistance_Association
Have_Toxicity_Association
be associated to
Each article was independently labeled by two annotators. discrepancies were resolved by a third reviewer.
d
e
The whole picture of GPAS knowledge graph
Relations from K. pneumoniae (degree = 137)

**Extended Data Fig. 12 Pathogen knowledge graph construction.** a, Workflow of pathogen knowledge graph construction. Comprehensive literature retrieval yielded 10,493 articles covering 1,242 pathogen species, enabling successful model fine-tuning. b, The complete GPAS knowledge graph visualization, displaying interconnected entities including pathogens, diseases, virulence factors, and clinical manifestations. c, Expanded view of relations centered on Klebsiella pneumoniae (degree = 137), illustrating the rich connectivity of the knowledge graph with associated diseases, virulence genes, resistance determinants, and literature evidence.

#### Extended Data Fig. 13 Big data for knowledge graph construction

**a**

Major Sample Types of Big Data

| Major Sample Type (Host) | Public | In House | Total |
|---|---|---|---|
| Digestive (Clinical Patient) | 6810 | 562 | 7372 |
| Respiratory (Clinical Patient) | 321 | 4217 | 4538 |
| Digestive (Wild Animal) | 1628 | 133 | 1761 |
| Tissue (Wild Animal) | 843 | 750 | 1593 |
| Soil Sediment (Environment) | 1446 | 29 | 1475 |
| Water (Environment) | 1395 | 66 | 1461 |
| Respiratory (Healthy People) | 0 | 1387 | 1387 |
| Body Fluid (Clinical Patient) | 0 | 848 | 848 |
| Respiratory (Wild Animal) | 787 | 0 | 787 |
| Other (Vector Animal) | 747 | 0 | 747 |
| Digestive (Domestic Animal) | 634 | 0 | 634 |
| Tissue (Vector Animal) | 0 | 469 | 469 |
| Atmosphere (Environment) | 310 | 62 | 372 |
| Environmental Other (Environment) | 118 | 55 | 173 |
| Tissue (Domestic Animal) | 127 | 0 | 127 |
| Body Fluid (Healthy People) | 0 | 106 | 106 |
| Body Fluid (Domestic Animal) | 0 | 88 | 88 |
| Earth Surface (Environment) | 60 | 0 | 60 |
| Digestive (Healthy People) | 0 | 46 | 46 |
| Other (Wild Animal) | 30 | 0 | 30 |
| Urogenital (Clinical Patient) | 0 | 28 | 28 |
| Body Surface (Clinical Patient) | 17 | 0 | 17 |
| Tissue (Clinical Patient) | 0 | 14 | 14 |
| Digestive (Vector Animal) | 12 | 0 | 12 |
| CNS (Clinical Patient) | 0 | 8 | 8 |
| Other (Clinical Patient) | 0 | 5 | 5 |
| Body Fluid (Wild Animal) | 0 | 4 | 4 |
| Other (Domestic Animal) | 2 | 0 | 2 |

**b**

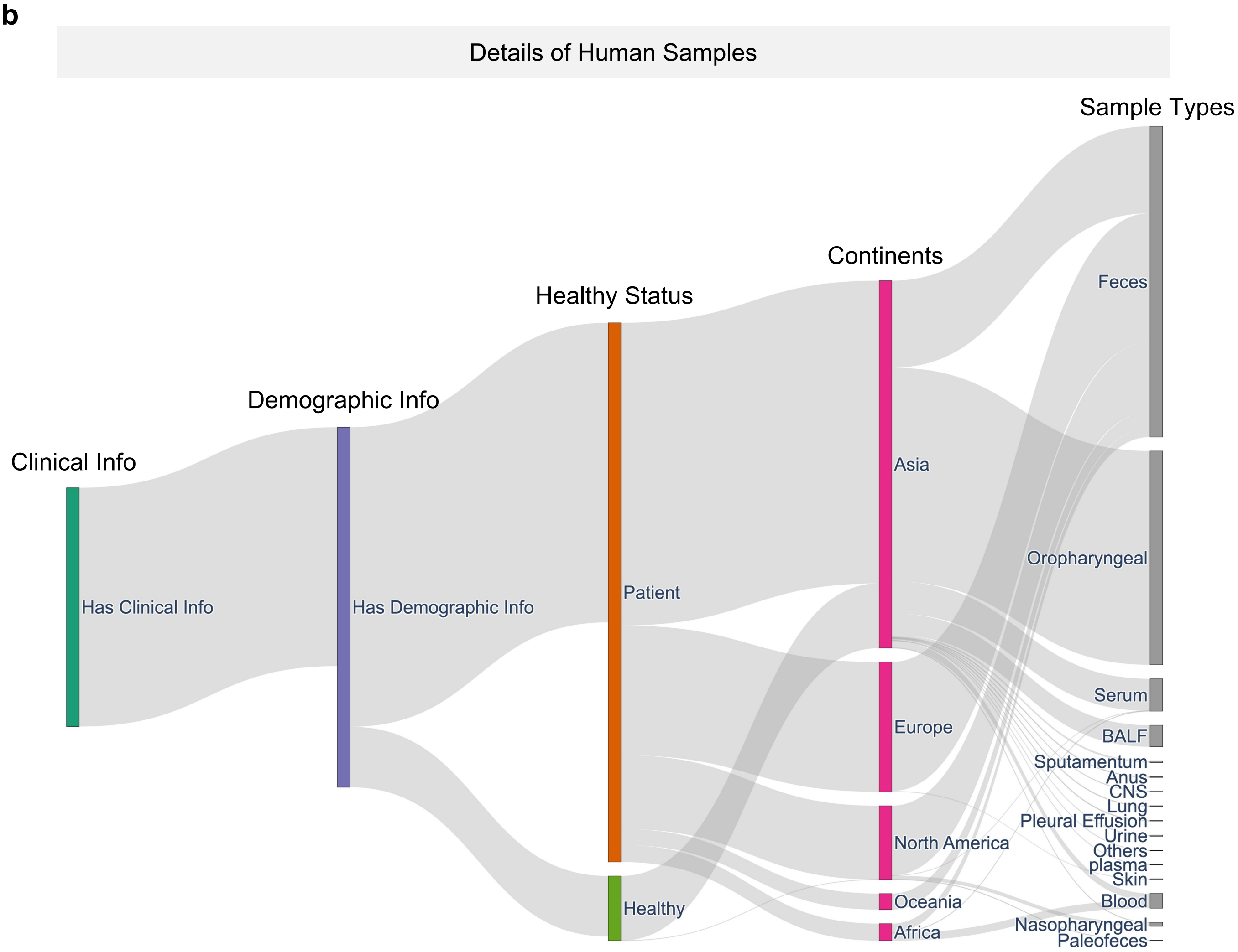

**Extended Data Fig. 13 Big data for knowledge graph construction.** a, Summary of major sample types and data sources incorporated into the GPAS knowledge graph. The table catalogs 24,164 samples by major sample type (Digestive, Respiratory, Tissue, Body Fluid, etc.), host category (Clinical Patient, Wild Animal, Environment, Healthy People, Vector Animal, Domestic Animal), and data source (Public and In House). Total sample counts are provided for each category. b, Geographic distribution and detailed sample types of human-derived specimens. The left panel illustrates the continental distribution of human samples (North America, Europe, Asia, Australia, South America, Africa, Oceania). The right panel enumerates specific human sample types incorporated in the knowledge graph, including feces, oropharyngeal swabs, nasopharyngeal swabs, pleural effusion, blood, cancer tissue, transplant specimens, anus swabs, CNS samples, lung tissue, urine, plasma, skin, and paleofeces, demonstrating the broad clinical and biological diversity captured in the GPAS big data infrastructure.

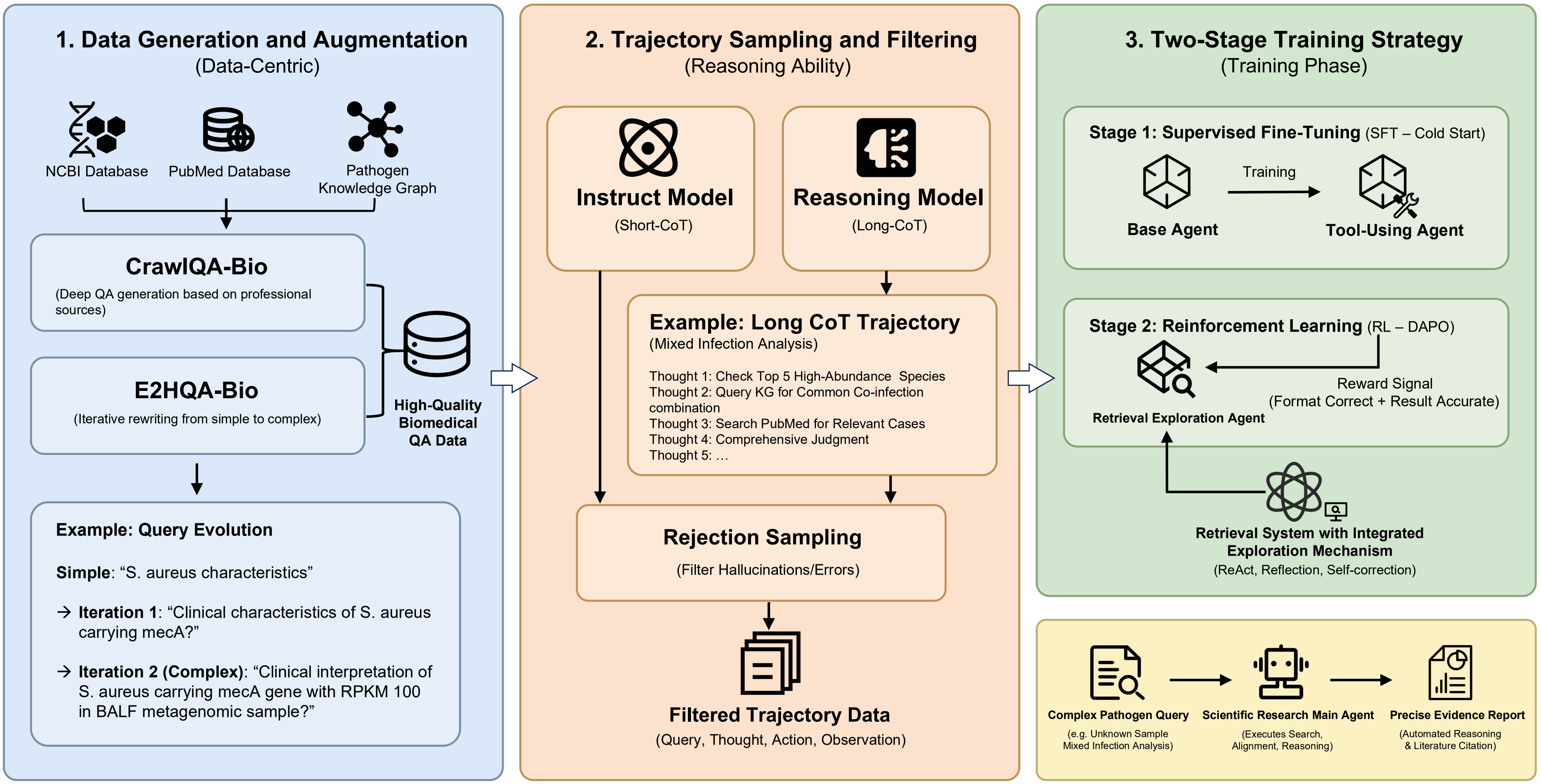
1. Data Generation and Augmentation
(Data-Centric)
NCBI Database
PubMed Database
Pathogen Knowledge Graph
CrawlQA-Bio
(Deep QA generation based on professional sources)
E2HQA-Bio
(Iterative rewriting from simple to complex)
High-Quality Biomedical QA Data
Example: Query Evolution
Simple: "S. aureus characteristics"
→ Iteration 1: "Clinical characteristics of S. aureus carrying mecA?"
→ Iteration 2 (Complex): "Clinical interpretation of S. aureus carrying mecA gene with RPKM 100 in BALF metagenomic sample?"
2. Trajectory Sampling and Filtering
(Reasoning Ability)
Instruct Model
(Short-CoT)
Reasoning Model
(Long-CoT)
Example: Long CoT Trajectory
(Mixed Infection Analysis)
Thought 1: Check Top 5 High-Abundance Species
Thought 2: Query KG for Common Co-infection combination
Thought 3: Search PubMed for Relevant Cases
Thought 4: Comprehensive Judgment
Thought 5: …
Rejection Sampling
(Filter Hallucinations/Errors)
Filtered Trajectory Data
(Query, Thought, Action, Observation)
3. Two-Stage Training Strategy
(Training Phase)
Stage 1: Supervised Fine-Tuning (SFT – Cold Start)
Base Agent
Training
Tool-Using Agent
Stage 2: Reinforcement Learning (RL – DAPO)
Retrieval Exploration Agent
Reward Signal
(Format Correct + Result Accurate)
Retrieval System with Integrated Exploration Mechanism
(ReAct, Reflection, Self-correction)
Complex Pathogen Query
(e.g. Unknown Sample Mixed Infection Analysis)
Scientific Research Main Agent
(Executes Search, Alignment, Reasoning)
Precise Evidence Report
(Automated Reasoning & Literature Citation)

**Extended Data Fig. 14 GPAS-LLM training pipeline.** Schematic overview of the three-stage GPAS-LLM training pipeline. Data Generation and Augmentation: High-quality biomedical QA pairs were generated from NCBI databases, public literature, and the pathogen knowledge graph, producing CrawlQA-Bio and E2HQA-Bio datasets. Trajectory Sampling and Filtering: Complex pathogen queries were iteratively refined through query evolution, generating long chain-of-thought (Long CoT) reasoning trajectories with ReAct, reflection, and self-correction mechanisms. Two-Stage Training Strategy: Stage 1 involved supervised fine-tuning (SFT) with cold start using both short and long CoT examples; Stage 2 employed reinforcement learning with DAPO (Dynamic Adaptive Policy Optimization) to filter hallucinations and errors, optimizing format correctness and result accuracy.

**a**

Performance comparison of GPAS-agent and DeepSeek models on fever sample diagnosis

| Sample type | diagnostic performance | GPAS-agent | DeepSeek |
|---|---|---|---|
| BALF | correctly diagnosed | 51 | 38 |
| Blood samples | | 3 | 0 |
| Lung tissue samples | | 0 | 0 |
| Oropharyngeal swab samples | | 1 | 2 |
| Pleural effusion samples | | 3 | 2 |
| Sputum samples | | 4 | 2 |
| BALF | Incorrectly diagnosed | 16 | 29 |
| Blood samples | | 0 | 3 |
| Lung tissue samples | | 1 | 1 |
| Oropharyngeal swab samples | | 1 | 0 |
| Pleural effusion samples | | 1 | 2 |
| Sputum samples | | 1 | 3 |
| Total | | 82 | 82 |

**b**

Number of confirmed pathogen cases in clinical samples

| Definitive pathogen identification | Total Number |
|---|---|
| *Acinetobacter baumannii* | 12 |
| *Staphylococcus aureus* | 8 |
| *Klebsiella pneumoniae* | 15 |
| *Mycobacterium tuberculosis* | 13 |
| *Aspergillus fumigatus* | 6 |
| *Stenotrophomonas maltophilia* | 5 |
| *Pseudomonas aeruginosa* | 12 |
| *Rhizopus* | 1 |
| *Legionella pneumophila* | 2 |
| *Burkholderia cepacia* | 1 |
| Severe acute respiratory syndrome coronavirus 2 | 4 |
| *Enterococcus faecium* | 3 |
| *Corynebacterium striatum* | 2 |
| *Aspergillus* | 12 |
| *Serratia marcescens* | 1 |
| *Pneumocystis jirovecii* | 1 |
| *Aspergillus flavus* | 1 |
| *Escherichia coli* | 5 |
| Influenza A virus | 1 |
| *Mycoplasmoides pneumoniae* | 2 |
| *Aspergillus terreus* | 1 |

**Extended Data Fig. 15 Dataset for diagnostic accuracy assessment.** a. Performance comparison of GPAS-agent and DeepSeek models on fever sample diagnosis. The table details sample numbers for the febrile patient cohort (n=100) spanning multiple sample types including sputum, blood, lung tissue, oropharyngeal swab, pleural effusion and bronchoalveolar lavage fluid. b, Number of confirmed pathogen cases in the clinically diagnosed cohort (n = 82).
